# Polygenic Scores of Core-1 Alzheimer’s Disease Biomarkers Predict Early Cognitive and Pathological Change

**DOI:** 10.1101/2025.07.12.25331438

**Authors:** Yuexuan Xu, Min Qiao, Tamil I. Gunasekaran, Yian Gu, Dolly Reyes-Dumeyer, Angel Piriz, Danurys Sanchez, Belisa Soriano, Yahaira Franco, Zoraida Dominguez Coronado, Patricia Recio, Diones Rivera Mejia, Martin Medrano, Rafael A. Lantigua, Lawrence Honig, Rachael Wilson, Jennifer J Manly, Adam M. Brickman, Corinne D. Engelman, Sterling Johnson, Sanjay Asthana, Badri Vardarajan, Richard Mayeux

**Author notes:** Correspondence: Richard Mayeux, MD, Gertrude H. Sergievsky Professor of Neurology, Psychiatry and Epidemiology Chair, Department of Neurology, and Neurologist-in-Chief, NewYork-Presbyterian/ Columbia University Irving Medical Center.

## Abstract

**BACKGROUND:** Core-1 biomarkers, such as amyloid PET, capture the earliest biological changes leading to Alzheimer’s disease (AD). While *APOE* is a major genetic factor, the contribution of other variants to Core-1 biomarkers remains unclear. The goal of this study is to determine whether genetic regulators of Core-1 biomarker levels predict AD pathology better than genetic regulators of clinical AD.

**METHODS:** Among 955 non-Hispanic white individuals, PGSs were built using GWAS of amyloid PET, plasma P-tau181, CSF P-tau181, and clinical AD. Hispanic-specific PGSs were constructed in 515 individuals using plasma P-tau181 and clinical AD GWAS. Baseline and longitudinal associations with plasma biomarkers and cognition were assessed, and replication was conducted in separate cohorts.

**RESULTS:** The Core-1 biomarker PGSs predicted AD pathology and associated cognitive performance better than the AD PGS in both populations.

**DISCUSSION:** The Core-1 PGS show improved predictive value for AD-related plasma biomarkers and early cognitive changes.

**HIGHLIGHTS:**

- *APOE*-ε4 explained more variance in plasma P-tau217 than in plasma P-tau181.
- PGSs based on Core-1 biomarkers outperformed AD PGSs in predicting plasma biomarkers and cognitive decline among asymptomatic individuals in white non-Hispanic and Hispanic individuals. However, the improvement in predictive power was modest and may vary by age.
- While the variance in P-tau181 and P-tau217 explained by individual Core-1 PGSs remains limited, the distinct genetic signals captured by the best-performing PGSs across different Core-1 biomarkers may provide an opportunity for developing an integrative Core-1 PGS that more effectively predicts plasma P-tau181 and P-tau217 levels than AD-based PGS.

**RESEARCH IN CONTEXT:** *SYSTEMATIC REVIEW:* Core-1 biomarkers, such as amyloid PET, capture the earliest biological changes in Alzheimer’s disease (AD). The authors reviewed the literature on the association between core-1 biomarker-based polygenic scores (PGS) and plasma biomarkers using traditional sources (e.g., PubMed), meeting abstracts, and presentations. While several studies examined the association between PGS and plasma P-tau181 and P-tau217, most PGSs were based on AD genetic studies and performed poorly in predicting these biomarkers—especially in asymptomatic individuals. No studies have comprehensively assessed Core-1 biomarker-based PGSs in relation to P-tau181, P-tau217, other plasma biomarkers, and cognitive change.

*INTERPRETATION:* Our findings show that Core-1 biomarker-based PGSs outperform AD PGSs in predicting early changes in plasma P-tau181, P-tau217, other non-tau plasma biomarkers, and cognition in asymptomatic White and Caribbean Hispanic individuals.

*FUTURE DIRECTIONS:* The Core-1 PGS offers improved predictive value for AD-related plasma biomarkers and early cognitive changes. Future research should validate these findings using larger genome-wide association studies and explore integrated Core-1 PGSs for P-tau217 and P-tau181, especially as more Core-1 biomarker genetic data become available.

## 1. BACKGROUND

Alzheimer’s disease (AD) is a progressive neurodegenerative disorder with a prolonged preclinical phase that may begin a decade or more before cognitive symptoms appear. Early detection and intervention are critical to slowing disease progression. To support this, the National Institute on Aging–Alzheimer’s Association (NIA-AA) Research Framework defines AD biologically, categorizing its biomarkers into two groups: Core-1, which reflects early amyloid and tau pathology, and Core-2, which provides prognostic information[1]. Among these, Core-1 biomarkers, such as amyloid PET imaging and cerebrospinal fluid (CSF) or plasma beta-amyloid (Aβ42) and phosphorylated tau (P-tau181, P-tau217, P-tau231), are central to the early detection of AD, even in asymptomatic individuals.

*APOE* is the primary genetic contributor to Core-1 biomarkers, but additional genetic predictors of early AD neuropathologic changes may be clinically relevant[2]. One promising approach involves using polygenic scores (PGSs), which capture the cumulative effects of common genetic variants and have been widely studied in relation to the clinical AD phenotype[3,4]. Several studies have examined associations between PGSs derived from genome wide association studies (GWASs) of the clinical AD phenotype and Core-1 biomarkers, but the findings have been mixed, particularly when *APOE* variants are excluded[5–7]. This suggests that clinical AD-based PGSs may not optimally capture the specific Core-1 biomarkers-related biological processes in the early stages of the disease. Variants identified by using the clinical AD phenotype GWASs may represent broader downstream disease mechanisms rather than those directly linked to early amyloid and tau pathology[8]. Supporting this idea, no association between an AD PGS and amyloid PET was observed using all AD-associated variants, but a significant association was found when restricting the analysis to variants also linked to amyloid burden[9].

Genetic investigations of AD have begun to focus on biomarker-specific pathologies, such as amyloid PET, CSF Aβ42, CSF P-tau181, and plasma P-tau181[7,10–15]. However, the predictive value of PGSs derived from these biomarkers, particularly in comparison to clinical AD PGSs or the *APOE* genotype, remains largely untested, especially in populations of different racial and ethnic groups. To address this gap, we sought to (1) derive *APOE*-independent PGSs based on GWASs of Core-1 AD biomarkers, including amyloid PET and P-tau181 from CSF and plasma, and to evaluate their utility in predicting plasma P-tau181/P-tau217 levels; (2) evaluate their ability to predict other downstream plasma biomarker changes and cognitive performance in non-demented older adults; and (3) assess their performance relative to *APOE* and GWAS-based clinical AD PGSs in both white non-Hispanic and Hispanic populations.

## 2. METHODS

### 2.1 Study participants

For this study, we collected data from five cohorts that enrolled non-Hispanic white and Caribbean Hispanic individuals. Data for non-Hispanic white participants came from the Alzheimer’s Disease Neuroimaging Initiative (ADNI), the Wisconsin Registry for Alzheimer’s Prevention (WRAP), and the Wisconsin Alzheimer’s Disease Research Center (Wisconsin-ADRC). Data for Caribbean Hispanic participants came from individuals who self-identified as Caribbean Hispanic enrolled in either the Estudio Familiar de Influencia Genética en Alzheimer (EFIGA) or the Washington Heights– Hamilton Heights–Inwood Columbia Aging Project (WHICAP). Details of the study design are described elsewhere (see Table 1 and Supplementary Methods)[16–20].

**Table 1.**
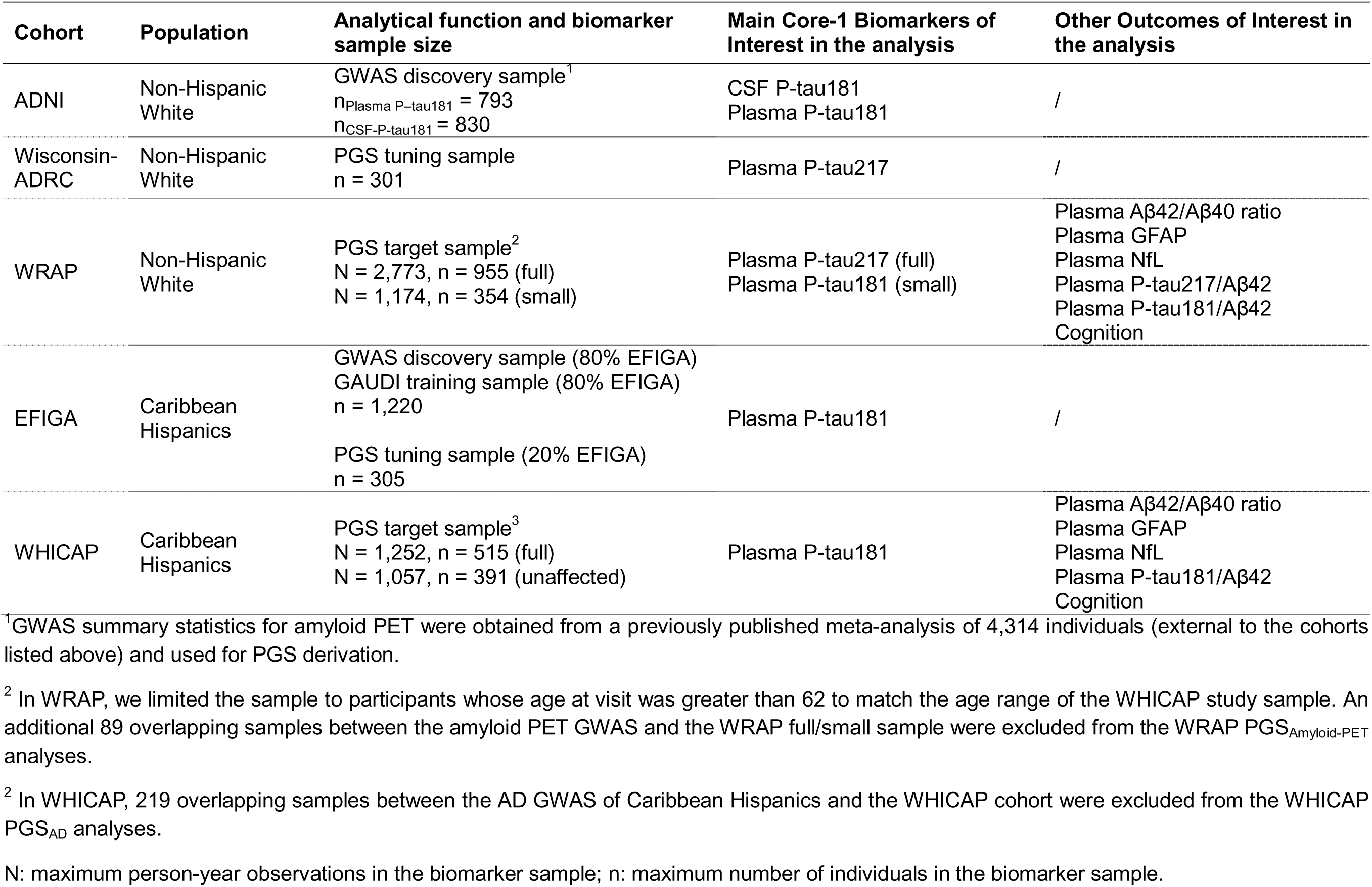
Overview of Analytical Samples.

Briefly, PGS calculation requires three datasets: (1) a base dataset with GWAS summary statistics (from public sources or in-house discovery if public data are unavailable), (2) a tuning dataset to optimize SNP linkage disequilibrium (LD) structure, and (3) a target dataset for PGS computation and downstream analyses. Among non-Hispanic white participants, we used ADNI as the GWAS discovery sample for Core-1 biomarkers to derive effect size estimates for SNPs. We used the Wisconsin-ADRC sample as the tuning sample, and WRAP data served as the target sample. Similarly, among Caribbean Hispanics, we divided the EFIGA data into 80% and 20% subsets: 80% were used as the GWAS discovery sample (for plasma P-tau181), and the remaining 20% were used as the PGS tuning sample. WHICAP data served as the target sample (Table 1).

WRAP and WHICAP are both longitudinal cohorts. Because the minimum enrollment age in WHICAP is 62 years, we limited the WRAP sample to participants whose age at interview was greater than 62 years to ensure comparability across cohorts. We treated the first interview after age 62 as the baseline in WRAP. All participants provided written informed consent prior to participation. The Columbia University Medical Center Institutional Review Board approved the study and conducted it in accordance with the Declaration of Helsinki.

### 2.2 Biomarker and neuropsychological assessments

We describe the details on the acquisition and processing of plasma and CSF samples from ADNI, WRAP, Wisconsin-ADRC, EFIGA, and WHICAP (see Supplementary Methods) [21–26]. We log_10_-transformed biomarker values due to skewed distributions. In the target samples, we examined the associations between PGSs for Core-1 biomarkers and a range of AD-related plasma biomarkers including plasma Aβ42/Aβ40 ratio, P-tau181, P-tau217, neurofilament light (NfL), glial fibrillary acidic protein (GFAP), P-tau181/Aβ42 ratio, and P-tau217/Aβ42 ratio.

We also analyzed the association of PGSs derived from Core-1 biomarkers with cognitive performance in the target samples. Previously, we described the details of cognitive assessments in WRAP and WHICAP [27,28]. Briefly, cognition in WRAP is assessed with a comprehensive neuropsychological battery, which includes domain-specific measures of immediate learning, delayed recall, and executive function, as well as the Preclinical Alzheimer’s Cognitive Composite 3 (PACC-3), a global cognition measure. In WHICAP, all participants completed a neuropsychological battery assessing episodic memory, language, visuospatial ability, and processing speed. In the current study, we used episodic memory, language, speed, and visuospatial composites derived from factor analysis as domain-specific outcome variables, and we used their average to derive a global cognitive functioning score. We used both domain-specific and global scores as outcomes in the current analysis.

### 2.3 DNA collection, genotyping, and quality control

We previously described the genomic data collection and quality control procedures for all study samples [28–30]. In brief, we processed genotyped data using standard quality control protocols in PLINK (v1.9) [31,32]. We removed single nucleotide polymorphisms (SNPs) if they had a minor allele frequency <1%, had a Hardy-Weinberg equilibrium test *P*-value <1×10⁻, or were missing in >5% of individuals. We excluded individuals with discrepancies between self-reported sex and genetically determined sex or with >5% missing genotype data. We then imputed genotypes using the Trans-Omics for Precision Medicine Imputation Reference Panel. In addition, we excluded post-imputation, SNPs with MAF <1%, or imputation quality scores (R²) <0.8. All data were aligned to the GRCh38 genome build. We first grouped the *APOE* genotype into six types based on rs7412 and rs429358, and then we combined them into two groups: ε4 carriers (ε2/ε4, ε3/ε4, ε4/ε4) and non-carriers (ε2/ε2, ε2/ε3, ε3/ε3).

To refine ancestry estimates while accounting for familial relatedness, we used the PC-relate method, a model-free approach [33]. We estimated the initial kinship coefficients using the KING algorithm implemented in the SNPRelate R package, based on a subset of autosomal SNPs pruned for LD using a 10 bp sliding window and an r² threshold of 0.1. We performed principal components analysis (PCA) with PC-AiR to capture the population structure while accounting for relatedness [34,35]. Ancestry inference was aided by projecting study samples onto reference PCs derived from 1,000 Genomes Project data. We used the first two PCs to adjust kinship estimates for genetic ancestry, which we then used to refine PCA. We reiterated this process twice, which resulted in a final set of ancestry PCs and a genetic relationship matrix adjusted for relatedness and population structure.

### 2.4 GWASs of Core-1 biomarkers and PGS calculation

The Core-1 biomarker availability varied across cohorts, thus we tailored GWASs, PGS tuning, and testing strategies accordingly. Details on the availability of Core-1 biomarkers used for GWAS discovery, PGS tuning and testing (CSF P-tau181, plasma P-tau181, and plasma P-tau217) are provided in Table 1.

In the current analyses among non-Hispanic white individuals, we derived PGSs for three Core-1 biomarkers, amyloid PET, CSF P-tau181, and plasma P-tau181, as well as an AD PGS based on a recent AD GWAS meta-analysis. We referred to these as PGSAmyloid-PET, PGSCSF-P-tau181, PGSPlasma-P-tau181, and PGSAD, respectively. We did not derive PGS for CSF and plasma Aβ42 due to their U-shaped relationship with age, which can introduce age-dependent and potentially misleading directions of SNP effect sizes [36,37]. PGS_Amyloid-PET_ was derived using summary statistics from a meta-analysis of amyloid PET imaging data from 4,314 participants previously conducted by our group [14]. To avoid overfitting bias, we excluded 89 individuals who overlapped between the amyloid PET GWAS meta-analysis sample and the WRAP cohort from the WRAP PGS_Amyloid-PET_ analyses. Although multiple GWASs have been conducted for CSF and plasma P-tau181, the summary statistics were either not publicly available or lacked essential information (e.g., beta estimates) required for PGS construction. For CSF P-tau181, two large meta-analyses have been published: Jansen et al. (2022)[13] and Deming et al. (2017)[15]. We did not use Jansen et al. because the meta-analysis employed a sample size–weighted approach and reported only z-scores, which precluded the extraction of beta estimates. Additionally, the inclusion of heterogeneous cohorts limited our ability to derive unbiased effect sizes. Although summary statistics - including beta estimates - from Deming et al. are available via NIAGADS (NG00055), they were inaccessible at the time of manuscript submission due to a platform transition. Similarly, no publicly available plasma P-tau181 GWAS provided the necessary information for PGS construction[10,12]. Therefore, we conducted GWASs for these two biomarkers using data from 830 participants with CSF and 793 participants with plasma P-tau181 measurements from ADNI. We derived PGS_AD_ from a recent meta-analysis of AD GWASs by Bellenguez et al [38].

Among Caribbean Hispanic individuals, we derived PGS_Plasma-P-tau181_ only for plasma P-tau181 and conducted a GWAS of plasma P-tau181 in 1,220 from the EFIGA cohort (approximately 80% of the full EFIGA cohort), given the limited availability of large-scale CSF and amyloid PET data for GWAS and PGS analyses. We also derived a PGS_AD_ based on a GWAS of case-control AD status among Caribbean Hispanic individuals conducted by Qiao et al [39]. We also excluded 219 individuals who overlapped between the AD GWAS sample and the WHICAP cohort from the WHICAP PGS_AD_ analyses. We conducted all GWAS analyses of CSF and plasma P-tau181 using PLINK v2.0, adjusting for age, sex, disease status, and the first 10 principal components (PC) to account for population stratification.

The primary method for calculating PGS was the clumping and thresholding (C+T) approach in both non-Hispanic white and Caribbean Hispanic individuals. Specifically, we computed each PGS using PLINK v2.0. We selected SNPs using LD clumping with a 1,000 KB window, an R^2^ threshold of 0.1, and a *P*-value threshold of 1.0. We excluded the *APOE* region (hg38 coordinates: chr19, 43,907,927 to 45,908,821). We performed PGS analyses for each available Core-1 biomarker using selected *P*-value thresholds ranging from 5e^-08^ to 1. We selected the *P*-value threshold that maximized prediction (e.g., maximum incremental R^2^) of Core-1 biomarkers in the tuning cohort (e.g., P-tau217 in Wisconsin-ADRC and P-tau181 in the EFIGA tuning sample) for application in the target sample.

Among Caribbean Hispanic individuals, we also calculated a GAUDI PGS for plasma P-tau181, referred to as PGS_GAUDI_ [40]. GAUDI is a powerful tool for PGS analyses in admixed populations, as it explicitly models ancestry-differential effects while borrowing information across segments with shared ancestry. We chose this approach because it showed improved performance over the C+T method and does not require external samples for GWAS discovery or parameter tuning, making it especially advantageous for underrepresented populations with limited sample size. Details of the GAUDI PGS methods are described elsewhere [40]. Briefly, we used the GWAS discovery sample of plasma P-tau181 as the training sample and limited variants to those with GWAS *P-* value < 0.05 for further variant selection and weight estimation. Local ancestry was inferred using RFMix and combined with haplotype dosage data to generate ancestry-specific genotype matrices [41]. We performed model training using GAUDI’s fused lasso framework with cross-validation to select the optimal *P*-value threshold. We applied the final GAUDI model to the target WHICAP sample to generate individual-level PGSs.

### 2.5 Statistical analysis

The baseline characteristics of the study population are presented as mean values with standard deviations for continuous variables and as percentages for categorical variables. We also assessed the correlations among the various genetic predictors of interest in this study separately in non-Hispanic white and Hispanic individuals.

We tested the associations of *APOE*-ε4, Core-1 biomarker PGSs, and the AD PGS with plasma biomarkers and cognition in WRAP for non-Hispanic white and in WHICAP for Hispanic individuals using available data for each outcome of interest. Among non-Hispanic white individuals, we focused on the predictive performance of the genetic predictors for plasma P-tau217 in the full WRAP sample and P-tau181 in a small subsample. We assessed the genetic association with P-tau217 in both full and small cohorts, whereas we assessed the association with P-tau181 in the smaller group only.

We further analyzed associations with P-tau217 and P-tau181 across age groups, sex, and *APOE*-ε4 allele status using longitudinal data. For age group analyses, we categorized participants based on their age of interview in the full sample as <70, 70–75, or >75 years old, and we grouped those in the smaller sample as <70 or >70 years old due to limited follow-up data at older ages and small sample size. In addition, we assessed the longitudinal associations of all genetic predictors with other plasma biomarkers of interest and cognition (Table 1) in both the full and smaller WRAP samples. We used likelihood ratio tests to examine significant interactions.

Among Hispanic individuals, we focused on the predictive performance of the genetic predictors for plasma P-tau181 only. Because WRAP includes only individuals free of dementia, to ensure comparability across ancestries, we tested the genetic associations with plasma P-tau181 in both the full WHICAP sample, and a subset restricted to dementia-free individuals. In both samples, we also evaluated PGS associations with P-tau181 across age groups, sex, and *APOE*-ε4 allele status using longitudinal data, applying the same stratification scheme used in WRAP, with the addition of an age >80 group in WHICAP due to its relatively older population. We also evaluated longitudinal associations of all genetic predictors with other plasma biomarkers of interest and cognition in the dementia-free WHICAP samples.

For baseline analyses, we used linear models with age, sex, and the first 10 PCs as covariates. For longitudinal biomarker analyses, we applied linear mixed-effects models with random intercepts to account for within-individual correlation due to repeated measurements, adjusting for age, sex, and the first 10 PCs. For longitudinal cognitive analyses, we additionally included practice effects (visit number – 1) as a covariate.

We evaluated the performance of the genetic predictors for the main Core-1 biomarkers of interest using *P*-values and incremental R² to compare the variance explained across PGS models. For mixed-effects models, we reported the incremental marginal coefficient of determination (incremental Nakagawa pseudo-R²). We considered the two-sided *P*-value < 0.05 statistically significant. Given the correlation among the outcomes, we did not apply formal multiple testing correction. Instead, we focused on consistent patterns across related biomarkers and reported all *P*-values to support interpretation in context. We also provided the effect sizes and confidence intervals to further contextualize the results. We z-standardized all continuous genetic predictors and conducted all statistical analyses using R v4.2.2.10.

### 2.6 Replication and sensitivity analysis

We conducted a series of internal replication and sensitivity analyses to assess the robustness of our main findings. Due to the limited availability of plasma biomarker and cognition data, we replicated key analyses in Wisconsin-ADRC, the non-Hispanic white WHICAP, and EFIGA cohorts.

Among non-Hispanic white individuals, we replicated both the longitudinal and stratified analyses of genetic associations with plasma P-tau217 across age groups, sex, and *APOE*-ε4 status using longitudinal data from cognitively normal participants in the Wisconsin-ADRC. We also replicated the longitudinal associations of the top-performing genetic predictors with other plasma biomarkers in the Wisconsin-ADRC. In WHICAP non-Hispanic white individuals, we replicated the longitudinal associations of these top-performing genetic predictors with cognition among non-demented participants. We also conducted sensitivity analyses on the predictive power of clinically informed AD PGSs on Core-1 biomarkers based on the largest clinical case-control GWAS meta-analysis by Kunkle et al[42]. In WRAP, plasma P-tau181 was collected earlier as part of a pilot study, resulting in some misalignment in age at assessment and participant IDs compared to P-tau217 and other biomarkers. Therefore, we re-estimated the associations of individual genetic predictors with both P-tau181 and P-tau217 using complete-case analysis.

Among Hispanic individuals, we performed internal replication analyses using the EFIGA tuning sample by testing associations between all genetic predictors and plasma biomarkers in both the full sample and the unaffected subset. These analyses also included evaluating the performance of PGS_GAUDI_ within the same cohort.

### 2.7 Integrative Core-1 biomarker PGS

We finally explored the integrative Core-1 biomarker PGS among non-Hispanic White individuals by summing the three Core-1 PGSs studied in this manuscript as an unweighted composite, referred to as PGS_Integrative_. All individual Core-1 PGSs were standardized before inclusion in the composite, and the final integrative PGS was also standardized to have a mean of 0 and a standard deviation of 1. We evaluated the association between PGS_Integrative_ and P-tau217, other plasma biomarkers, and cognition in the WRAP cohort, with replication of biomarker prediction in Wisconsin-ADRC and cognition in WHICAP non-Hispanic white cohort. We also performed stratified analyses by age group, sex, and *APOE* allele status.

## 3. RESULTS

### 3.1 Participant characteristics

The descriptive statistics of the participants from WRAP and WHICAP are in Table 2. Briefly, among non-Hispanic white individuals, after applying exclusion criteria, a total of 955 individuals remained in the cohort. The mean age at baseline was 65.71 years, the mean education level was 15.91 years, approximately 32% of WRAP participants were men, and about 39% were *APOE-*ε4 carriers. The mean number of visits was 2.91.

**Table 2.** Descriptive Statistics of the Target Biomarker Sample Used in the Analysis.

| Characteristic | Non-Hispanic Whites |  | p-value<sup>2</sup> | Caribbean Hispanics |
| --- | --- | --- | --- | --- |
|  | WRAP Full<br>N = 955<sup>1</sup> | WRAP Small<sup>a</sup><br>N = 354<sup>1</sup> |  | WHICAP<br>N = 515<sup>1</sup> |
| Age (years) | 65.71 (3.22) | 65.48 (3.17) | 0.2 | 75.35 (6.20) |
| **Sex** |  |  | 0.5 |  |
| Men | 304 (32%) | 121 (34%) |  | 146 (28%) |
| Women | 651 (68%) | 233 (66%) |  | 369 (72%) |
| Education (years) | 15.91 (2.27) | 16.09 (2.27) | 0.2 | 7.44 (4.49) |
| Maximum visits<sup>b</sup> | 2.91 (1.44) | 3.32 (1.45) | <0.001 | 2.44 (0.83) |
| AD | / | / |  | 115 (23%) |
| **APOE** |  |  | 0.5 |  |
| 2\2 | 3 (0%) | 0 (0%) |  | 4 (1%) |
| 2\3 | 82 (9%) | 40 (11%) |  | 76 (15%) |
| 2\4 | 30 (3%) | 12 (3%) |  | 9 (2%) |
| 3\3 | 501 (52%) | 173 (49%) |  | 323 (63%) |
| 3\4 | 302 (32%) | 117 (33%) |  | 98 (19%) |
| 4\4 | 37 (4%) | 12 (3%) |  | 4 (1%) |
| P-tau181 (pg/mL) | / | 2.69 (1.33) |  | 2.87 (3.75) |
| P-tau217 (pg/mL) | 0.39 (0.25) | 0.38 (0.24) | 0.7 | / |
| Aβ42/Aβ40 | 0.07 (0.02) | 0.07 (0.01) | >0.9 | 0.05 (0.06) |
| P-tau181/Aβ42 | / | 0.46 (0.28) |  | 0.50 (1.46) |
| P-tau217/Aβ42 | 0.07 (0.06) | 0.07 (0.05) | 0.6 | / |
| GFAP (pg/mL) | 120.98 (61.33) | 120.99 (62.41) | >0.9 | 215.48 (560.67) |
| NfL (pg/mL) | 17.19 (12.48) | 16.40 (7.87) | 0.2 | 24.73 (16.48) |

<sup>1</sup> Mean (SD); n (%)
<sup>2</sup> Welch Two Sample t-test; Pearson's Chi-squared test
<sup>a</sup> The WRAP small sample refers to a subsample in which plasma P-tau181 is available.
<sup>b</sup> Maximum visits reflect the longest follow-up available for each participant in the biomarker sample, which may vary across different outcomes.

Using the same inclusion criteria, a smaller sample (with plasma P-tau181 available) included a total of 354 individuals. There were no statistically significant differences in baseline characteristics or plasma biomarker levels between participants in the full WRAP sample and those in the smaller sample, except for the maximum number of follow-up visits.

515 Hispanic individuals from WHICAP were included in the analyses, with a maximum number of visits of 2.44. Compared to WRAP participants, WHICAP participants were older and had less education, with a mean baseline age of 75.35 years and a mean of 7.44 years of education. Approximately 28% of WHICAP participants were men, 22% were *APOE-*ε4 carriers, and 23% eventually developed dementia. Demographics related to the GWAS discovery sample and the PGS tuning sample are presented in Supplementary Table 1.

Among non-Hispanic white individuals, there were only negligible (ρ < 0.1) pairwise correlations observed among the genetic predictors. In Hispanic individuals, there were negligible correlations between the AD PGS and either *APOE*-ε4, PGS_GAUDI_ or PGS_Plasma-P-tau181_, although a moderate correlation was observed between PGS_GAUDI_ and PGS_Plasma-P-tau181_ (Supplementary Figure 1).

### 3.2 Baseline and longitudinal associations of APOE-ε4, AD PGS, and Core-1 biomarker PGSs with plasma P-tau217 and P-tau181 among non-Hispanic white individuals

*APOE*-ε4 was significantly associated with baseline plasma P-tau217 levels (β = 0.131, 95% CI: 0.101–0.161), explaining ∼7.9% of the variance. We found significant associations of baseline P-tau217 for PGS_Amyloid-PET_ (β = 0.020, 95% CI: 0.002–0.038) and PGS_CSF-P-tau181_ (β = 0.021, 95% CI: 0.006–0.036), as well as marginal associations for PGS_Plasma-P-tau181_ (β = 0.014, 95% CI: –0.001–0.030), though all explained substantially less variance (ΔR² < 0.01). PGS_AD_ was not associated with P-tau217 (Figure 1A).

**Figure 1.**
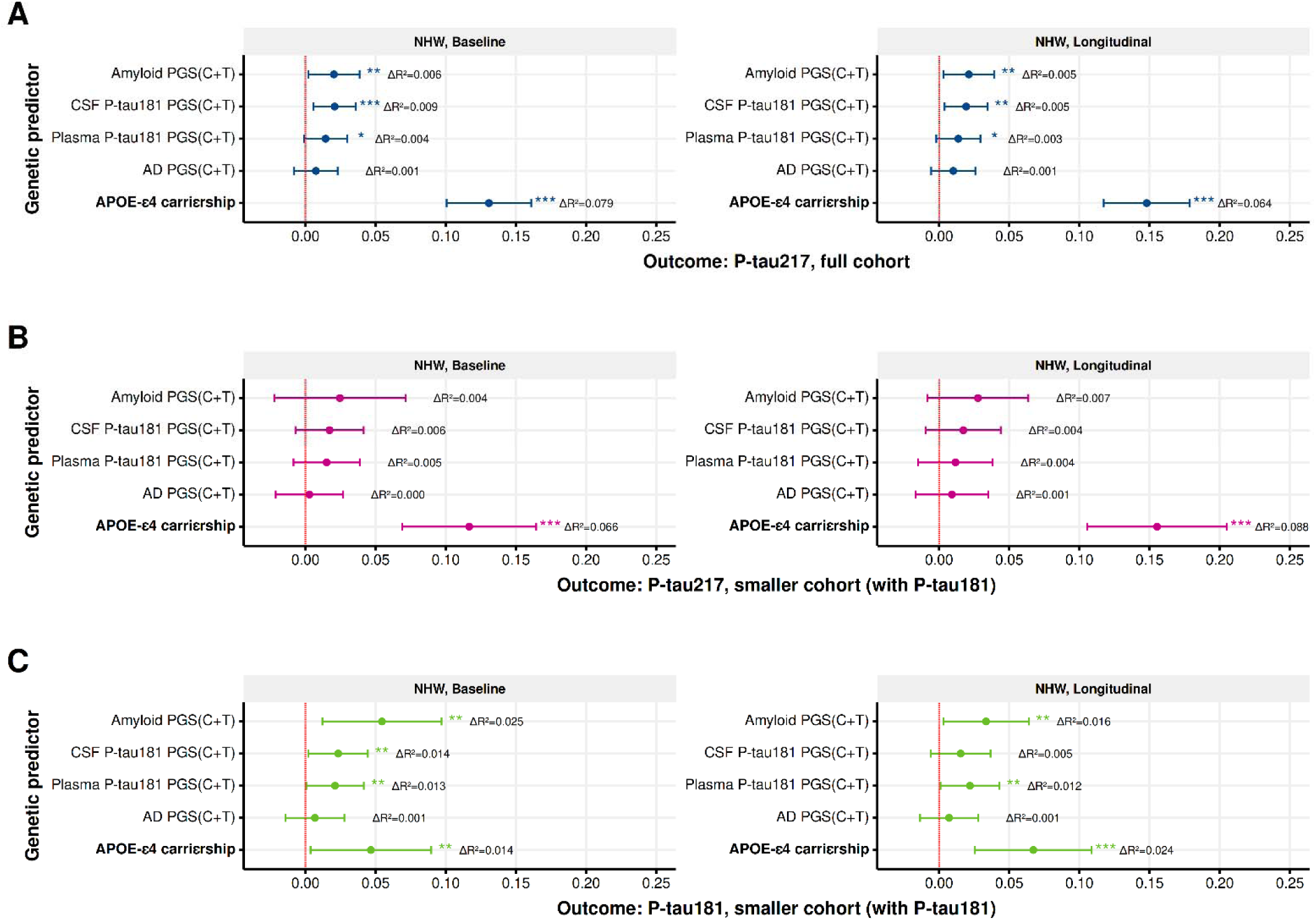
Associations of *APOE* ε4, Core-1 Biomarker-Derived PGSs, and AD GWAS PGS with Plasma P-tau181 and P-tau217 Among Non-Hispanic White Individuals (NHW full cohort: n_max_ = 955, N_max_ = 2,773; NHW smaller cohort: n_max_ = 354, N_max_ = 1,174) Figure 1 presents the baseline and longitudinal associations and predictive performance of *APOE* ε4, Core-1 biomarker-derived PGSs, and the AD GWAS PGS with plasma P-tau181 and P-tau217 among non-Hispanic White individuals using WRAP data. Since plasma P-tau217 is available in the full sample and P-tau181 is only available in a subset, we conducted three analyses. In Panel A, we show the association and predictive performance of all genetic predictors with plasma P-tau217 in the full sample. In Panel B, we show the same metrics for plasma P-tau217 in the subsample where plasma P-tau181 is also available. In Panel C, we present the associations and predictive performance for all genetic predictors with plasma P-tau181. All PGS predictors were first tuned in Wisconsin-ADRC, and the PGSs that showed the best prediction for P-tau217 in Wisconsin-ADRC were applied to the WRAP sample. To make the results comparable to the Caribbean Hispanic sample, we restricted WRAP participants to those aged 62 and older. The first interview after age 62 was treated as the baseline in WRAP. For baseline analyses, we used linear models adjusted for age, sex, and the first 10 principal components. For longitudinal analyses, we used linear mixed-effects models with the same covariates, and included random intercepts to account for within-individual correlations. ΔR² represents the incremental R², calculated as the difference in adjusted R² between models with and without the genetic predictor of interest. For linear mixed-effects models, ΔR² is calculated as the difference in Nakagawa’s R² with and without the genetic predictor. For the analyses related to the PGS derived from amyloid PET imaging, we additionally excluded 89 WRAP participants who were also included in the GWAS. All continuous genetic predictors were z-standardized. ***p < 0.01, **p < 0.05, *p < 0.1; C+T: clumping and thresholding; NHW: Non-Hispanic Whites; n_max_: Maximum number of unique individuals included in the sample. Actual sample size varies by outcome/predictor due to exclusion of individuals who were also part of the discovery GWAS. N_max_: Maximum number of person-level observations (i.e., person-years). The number of observations varies by outcome due to missing biomarker data at one or more follow-up visits.

We observed similar patterns for longitudinal associations. *APOE*-ε4 remained significantly associated with plasma P-tau217 over time (β = 0.148, 95% CI: 0.117– 0.179, ΔR² = 0.064). We again observed significant associations for PGS_Amyloid-PET_ (β = 0.021, 95% CI: 0.003–0.039) and PGS_CSF-P-tau181_ (β = 0.019, 95% CI: 0.004–0.035), as well as marginal associations for PGS_Plasma-P-tau181_ (β = 0.014, 95% CI: –0.002–0.029); all explained <1% of the variance. PGS_AD_ was not associated with longitudinal P-tau217 (Figure 1A).

Among the non-Hispanic white individuals (with plasma P-tau181 available), *APOE*-ε4 was significantly associated with P-tau217 levels at both baseline (β = 0.117, 95% CI: 0.069–0.165, ΔR² = 0.066) and longitudinally (β = 0.155, 95% CI: 0.106–0.205, ΔR² = 0.088). No PGSs were significantly associated with P-tau217 in this group, though effect sizes were consistent with the full cohort (Figure 1B).

For the non-Hispanic white individuals with P-tau181 as the outcome, *APOE*-ε4 was significantly associated at baseline (β = 0.047, 95% CI: 0.004–0.090, ΔR² = 0.014). Significant associations were also found for PGS_Amyloid-PET_ (β = 0.055, 95% CI: 0.012– 0.097, ΔR² = 0.025), PGS_CSF-P-tau181_ (β = 0.023, 95% CI: 0.002–0.045, ΔR² = 0.014), and PGS_Plasma-P-tau181_ (β = 0.021, 95% CI: 0.000–0.042, ΔR² = 0.013). PGS_AD_ was not associated. Longitudinally, *APOE*-ε4 remained significantly associated with plasma P-tau181 (β = 0.067, 95% CI: 0.026–0.109, ΔR² = 0.024). Significant associations were also observed for PGS_Amyloid-PET_ (β = 0.033, 95% CI: 0.003–0.064, ΔR² = 0.016) and PGS_Plasma-P-tau181_ (β = 0.022, 95% CI: 0.001–0.043, ΔR² = 0.012), whereas PGS_CSF-P-tau181_ and PGS_AD_ showed no associations (Figure 1C).

### 3.3 Baseline and longitudinal associations of APOE-ε4, AD PGS, and Core-1 biomarker PGSs with plasma P-tau181 among Hispanic individuals

In the Hispanic cohort (Figure 2A), only PGS_GAUDI_ was significantly associated with baseline plasma P-tau181 levels (β = 0.038, 95% CI: 0.013–0.061, ΔR² = 0.016). Longitudinally, plasma P-tau181 was associated with both *APOE*-ε4 (β = 0.049, 95% CI: 0.001–0.096, ΔR² = 0.005) and PGS_GAUDI_ (β = 0.026, 95% CI: 0.006–0.046, ΔR² = 0.009).

**Figure 2.**
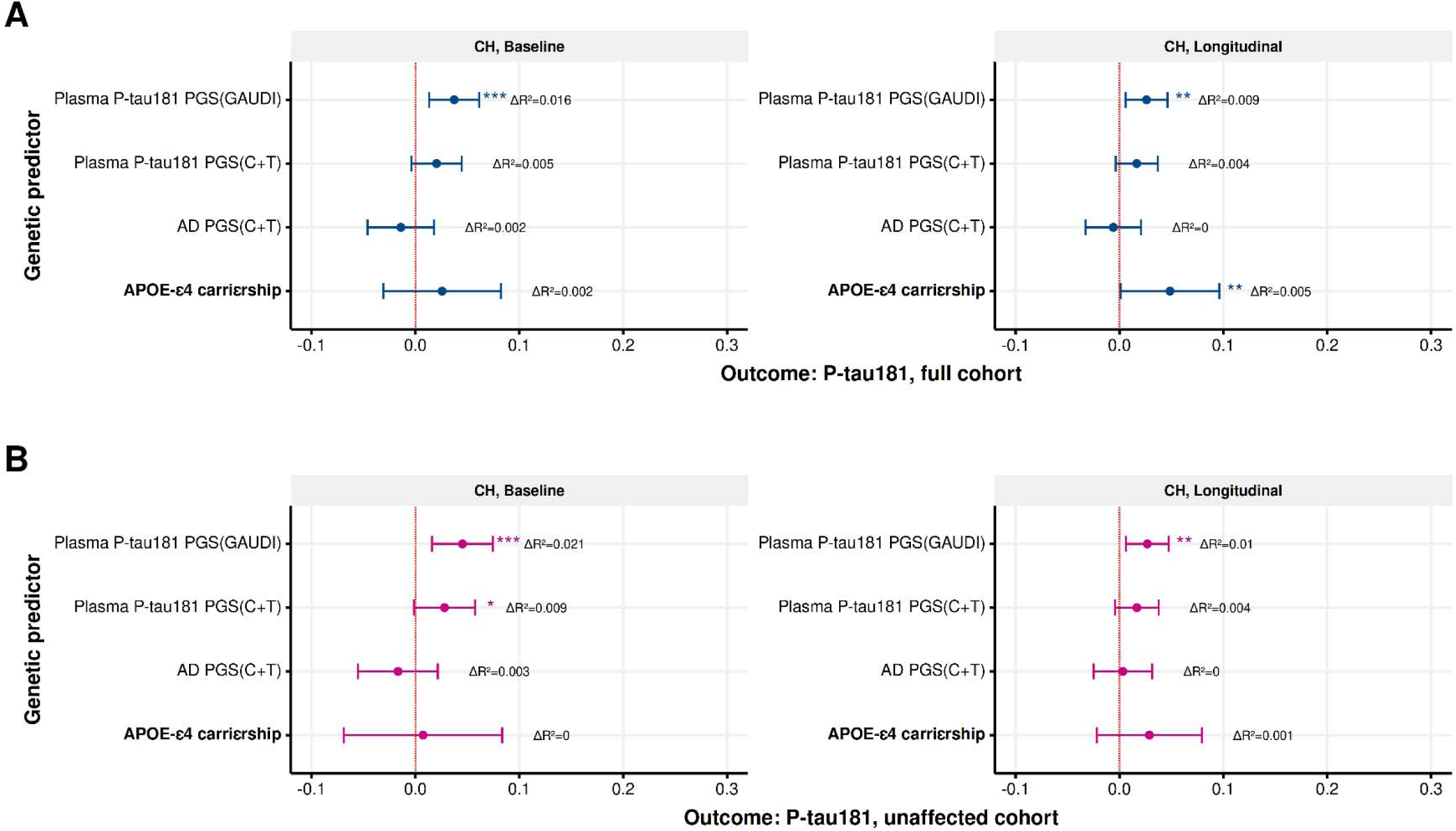
Associations of *APOE* ε4, Core-1 Biomarker-Derived PGSs, and AD GWAS PGS with Plasma P-tau181 Among Caribbean Hispanics Individuals (CH full cohort: n_max_ = 515, N_max_ = 1,252; CH unaffected cohort: n_max_ = 391, N_max_ = 1,057) Figure 2 presents the baseline and longitudinal associations and predictive performance of *APOE* ε4, Core-1 biomarker-derived PGSs, and the AD GWAS PGS with plasma P-tau181 among Caribbean Hispanic individuals using WHICAP data. Since WRAP includes only asymptomatic individuals while WHICAP includes both demented and non-demented individuals, we conducted two analyses to allow for comparison across ancestries. In Panel A, we show the association and predictive performance of all genetic predictors with plasma P-tau181 in the full WHICAP sample. In Panel B, we show the same metrics for the subsample of non-demented individuals only. All PGS predictors were first tuned in a tuning sample selected from EFIGA, and the PGSs that showed the best prediction for P-tau181 in the EFIGA tuning sample were applied to the WHICAP sample. For baseline analyses, we used linear models adjusted for age, sex, and the first 10 principal components. For longitudinal analyses, we used linear mixed-effects models with the same covariates, and included random intercepts for subjects to account for within-individual correlations. ΔR² represents the incremental R², calculated as the difference in adjusted R² between models with and without the genetic predictor of interest. For linear mixed-effects models, ΔR² was calculated as the difference in Nakagawa’s R² with and without the genetic predictor. For analyses involving the PGS derived from AD GWAS in the Hispanic sample, we excluded 219 WHICAP participants who were also included in the original GWAS to avoid sample overlap. All continuous genetic predictors were z-standardized. ***p < 0.01, **p < 0.05, *p < 0.1; C+T: clumping and thresholding; GAUDI: a powerful tool for PGS analyses in admixed populations, as it explicitly models ancestry-differential effects while borrowing information across segments with shared ancestry; CH: Caribbean Hispanics; n_max_: Maximum number of unique individuals included in the sample. Actual sample size varies by predictor due to exclusion of individuals who were also part of the discovery GWAS. N_max_: Maximum number of person-level observations (i.e., person-years).

Among non-demented Hispanic individuals (Figure 2B), PGS_GAUDI_ remained significantly associated with baseline plasma P-tau181 (β = 0.045, 95% CI: 0.016–0.075, ΔR² = 0.021), and a marginal association was observed for PGS_Plasma-P-tau181_ (β = 0.028, 95% CI: –0.001–0.057, ΔR² = 0.009). Longitudinally, only PGS_GAUDI_ was significantly associated with plasma P-tau181 (β = 0.027, 95% CI: 0.006–0.047, ΔR² = 0.010).

### 3.4 Stratified analyses

Using longitudinal WHICAP and WRAP data, we conducted stratified analyses by age group, sex, and *APOE*-ε4 carrier status. Among non-Hispanic white individuals, *APOE*-ε4 was associated with plasma P-tau217 across all age groups, with effect sizes increasing with age. For PGS_AD_, PGS_Plasma-P-tau181_, and PGS_CSF-P-tau181_, we only observed significant associations with plasma P-tau217 in the younger group (e.g., <70 years). In contrast, PGS_Amyloid-PET_ showed significant associations only in individuals aged 70 and older. Using a likelihood ratio test, we only observed a statistically significant interaction between age and *APOE-*ε4 (Supplementary Figure 2).

In the smaller cohort where plasma P-tau217 was the outcome, *APOE*-ε4 was associated with plasma P-tau217 across all age groups, and PGS_Amyloid-PET_ showed significant associations only in individuals aged 70 and older. However, no associations were observed with PGS_AD_, PGS_Plasma-P-tau181_, or PGS_CSF-P-tau181_ at any age group. The likelihood ratio test showed significant interactions between age and both *APOE*-ε4 and PGSAmyloid-PET.

For the smaller group where plasma P-tau181 was the outcome, *APOE*-ε4 was associated with plasma P-tau181 across all age groups. Significant associations were also observed with PGS_Plasma-P-tau181_ in individuals younger than 70, and PGS_Amyloid-PET_ continued to show significant associations only in individuals aged 70 and older. Likelihood ratio tests showed no significant interactions.

Among Hispanic individuals and the entire cohort of individuals, *APOE-*ε4 was associated with plasma P-tau181 in the older age groups (e.g., age >75 years). PGS_GAUDI_ and PGS_Plasma-P-tau181_ showed associations in the younger group (e.g., age <70 years). Similar findings were observed in the unaffected cohort, except for the fact that *APOE*-ε4 showed no association in any age group. In the total and unaffected groups, we only observed a statistically significant interaction identified by the likelihood ratio test between age and PGS_Plasma-P-tau181_.

Sex- and *APOE*-ε4-stratified analyses are shown in Supplementary Figure 3. We did not observe significant interactions between any genetic predictors and sex. However, the likelihood ratio test revealed a significant interaction between *APOE*-ε4 and PGS_AD_ in relation to P-tau217 in the total and smaller sample, with stronger PGS_AD_ effects observed among *APOE*-ε4 carriers.

### 3.5 Predictive power of Core-1 PGS for cognition and other plasma biomarkers among asymptomatic individuals

We assessed the longitudinal associations between PGSs of Core-1 biomarkers and other biomarkers beyond plasma P-tau181 and P-tau217 among asymptomatic individuals (Table 3). In non-Hispanic white individuals, *APOE*-ε4 was significantly associated with plasma Aβ42/Aβ40, GFAP, and P-tau217/Aβ42. PGS_Amyloid-PET_ was associated with Aβ42/Aβ40 and P-tau217/Aβ42. Both PGS_CSF-P-tau181_ and PGS_AD_ were associated with P-tau217/Aβ42.

**Table 3.**
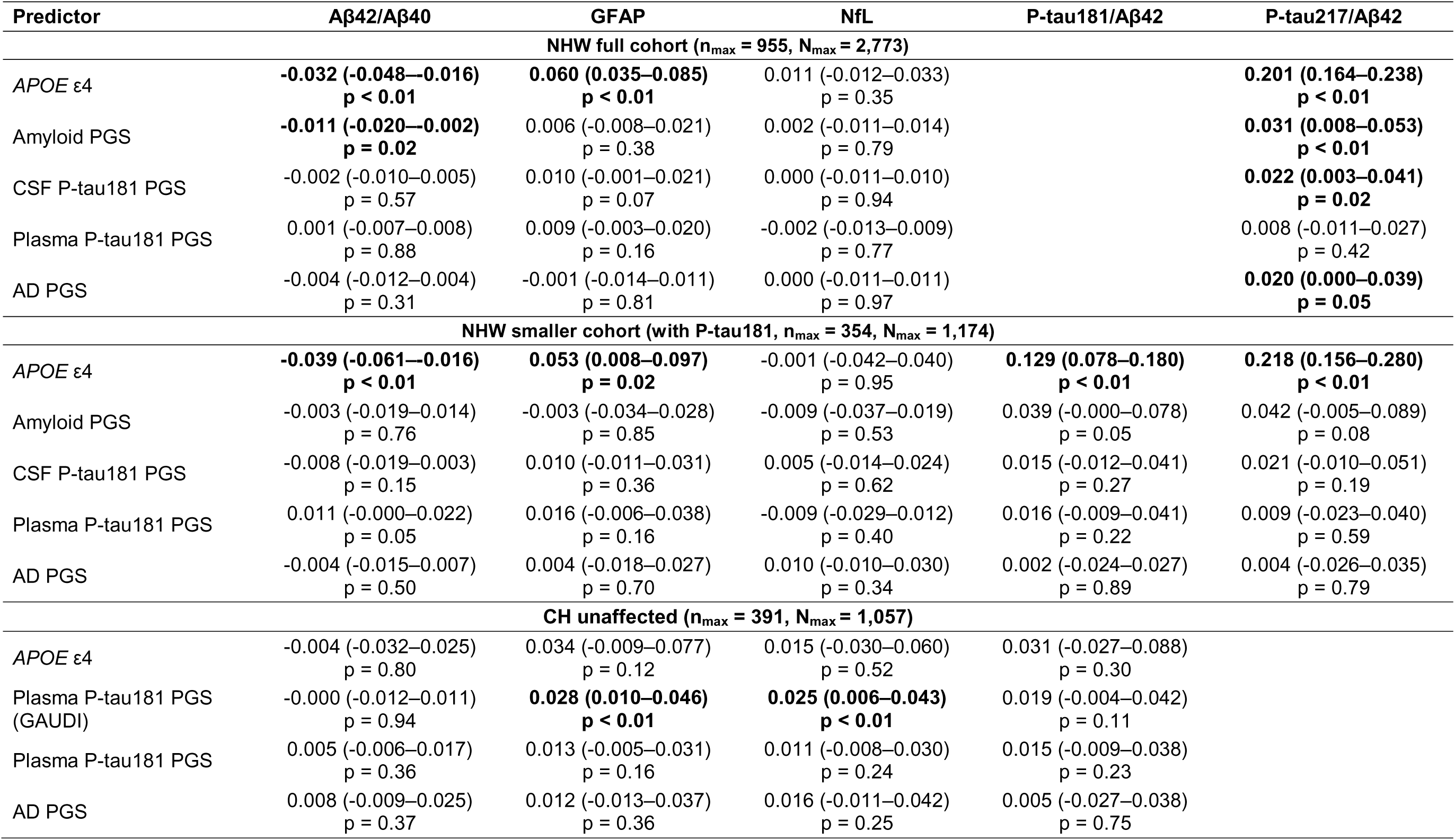
Associations of Genetic Predictors with Longitudinal Plasma Biomarkers Among Non-Demented Individuals. Table 3 presents the longitudinal associations of *APOE* ε4, Core-1 biomarker-derived PGSs, and the AD GWAS PGS with plasma biomarkers—beyond plasma P-tau181 and P-tau217—among non-Hispanic White and Caribbean Hispanic individuals who are non-demented. Among non-Hispanic Whites, we conducted two sets of analyses: one in the full WRAP sample and another in a subsample with available P-tau181 data. For Caribbean Hispanics, we tested longitudinal associations using only non-demented individuals with available longitudinal data. To ensure comparability with the Caribbean Hispanic sample, WRAP participants were restricted to those aged 62 and older. For all longitudinal analyses, we used linear mixed-effects models adjusted for age, sex, and the first 10 principal components, and included random intercepts for subjects to account for within-individual correlations. For analyses involving the PGS derived from amyloid PET imaging in the non-Hispanic white cohort, we excluded 89 WRAP participants who were also included in the original GWAS to avoid sample overlap. For analyses involving the PGS derived from AD GWAS in the Hispanic sample, we excluded 219 WHICAP participants who were also included in the original GWAS to avoid sample overlap. All continuous genetic predictors were z-standardized. 95% confidence intervals are shown in parentheses, and bolded values indicate p < 0.05. C+T: clumping and thresholding; GAUDI: a powerful tool for PGS analysis in admixed populations, as it explicitly models ancestry-differential effects while borrowing information across segments with shared ancestry; NHW: Non-Hispanic Whites; CH: Caribbean Hispanics; n_max_: Maximum number of unique individuals included in the sample. Actual sample size varies by outcome/predictor due to exclusion of individuals who were also part of the discovery GWAS. N_max_: Maximum number of person-level observations (i.e., person-years). The number of observations varies by outcome due to missing biomarker data at one or more follow-up visits.

In the smaller cohort of non-Hispanic white individuals, *APOE*-ε4 was associated with plasma Aβ42/Aβ40, GFAP, P-tau181/Aβ42, and P-tau217/Aβ42, whereas PGS_Amyloid-PET_ was marginally associated with P-tau181/Aβ42 and P-tau217/Aβ42. Among Hispanic individuals, only PGS_GAUDI_ was associated with GFAP and NfL.

For longitudinal cognition (Table 4), in non-Hispanic white individuals, *APOE*-ε4 and PGS_Amyloid-PET_ were associated with all domain-specific and global cognition measures. In the smaller group, only PGS_Plasma-P-tau181_ showed a significant association with executive function. Among Hispanic individuals, only PGS_Plasma-P-tau181_ showed an association with global cognition and processing speed.

**Table 4.**
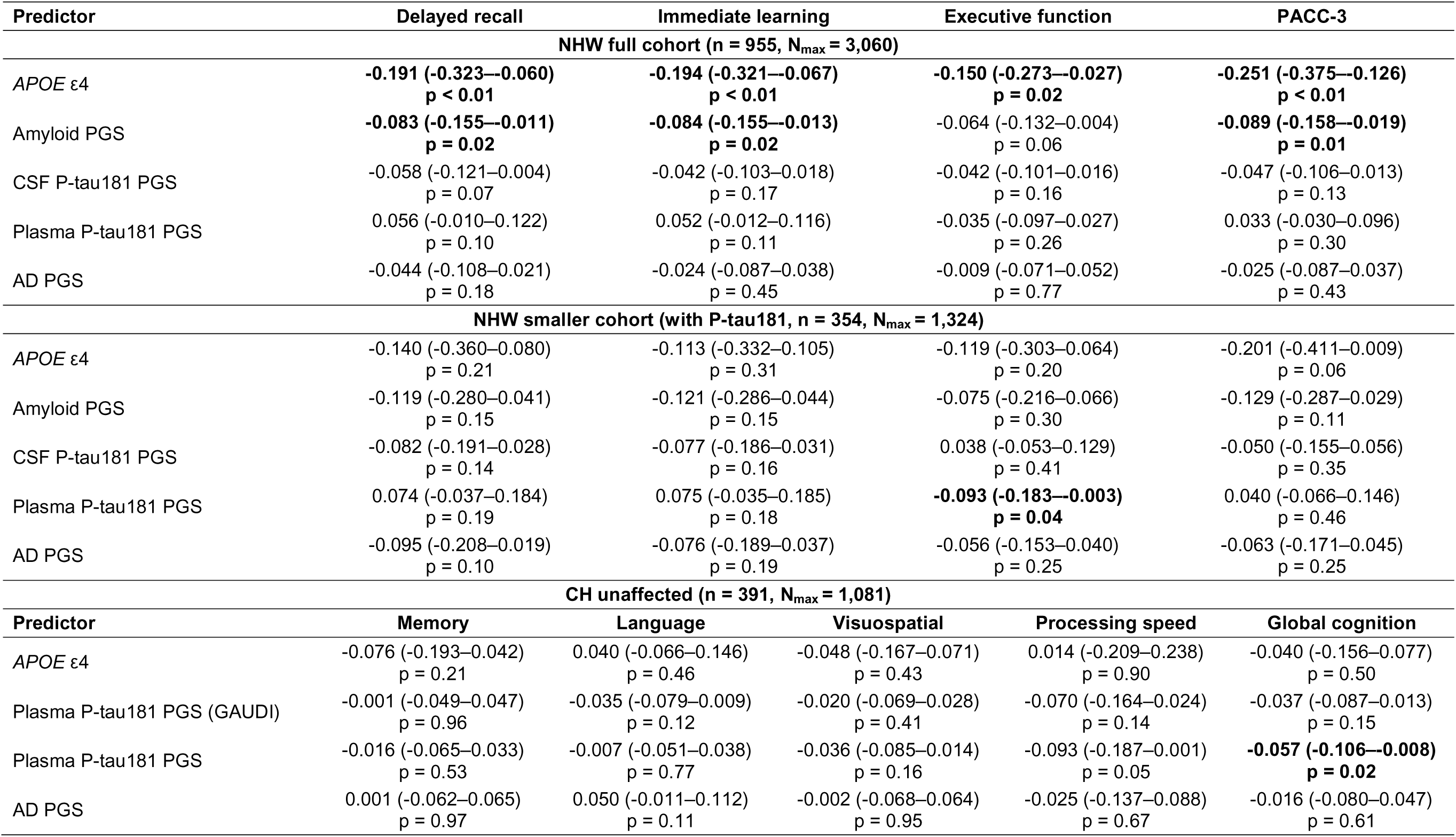
Associations of Genetic Predictors with Longitudinal Cognition Among Non-Demented Individuals. Table 4 presents the longitudinal associations of *APOE* ε4, Core-1 biomarker-derived PGSs, and the AD GWAS PGS with longitudinal cognitive outcomes among non-Hispanic White and Caribbean Hispanic individuals who are non-demented. Among non-Hispanic Whites, we conducted two sets of analyses: one in the full WRAP sample and another in a subsample with available P-tau181 data. For Caribbean Hispanics, we tested longitudinal associations using only non-demented individuals with available follow-up data. To ensure comparability with the Caribbean Hispanic sample, WRAP participants were restricted to those aged 62 and older. For all longitudinal analyses, we used linear mixed-effects models adjusted for age, sex, practice effects (visit number minus one), and the first 10 principal components, and included random intercepts for subjects to account for within-individual correlations. For analyses involving the PGS derived from amyloid PET imaging in the non-Hispanic white cohort, we excluded 89 WRAP participants who were also included in the original GWAS to avoid sample overlap. For analyses involving the PGS derived from AD GWAS in the Hispanic sample, we excluded 219 WHICAP participants who were also included in the original GWAS to avoid sample overlap. All continuous genetic predictors were z-standardized. 95% confidence intervals are shown in parentheses, and bolded values indicate p < 0.05. C+T: clumping and thresholding; GAUDI: a powerful tool for PGS analysis in admixed populations, as it explicitly models ancestry-differential effects while borrowing information across segments with shared ancestry; NHW: Non-Hispanic Whites; CH: Caribbean Hispanics. n_max_: Maximum number of unique individuals included in the sample. The actual sample size varies by outcome/predictor due to exclusion of individuals who were part of the discovery GWAS and because some cognitive assessments were not administered to all participants. N_max_: Maximum number of person-level observations (i.e., person-years). The number of observations varies by outcome due to missing cognitive data at one or more follow-up visits.

### 3.6 Replication and sensitivity analyses

We conducted several replication and sensitivity analyses to assess the robustness of our results. First, we replicated all biomarker analyses using longitudinal data from non-demented individuals in the Wisconsin-ADRC cohort (N = 247, n = 299). Similar to the WRAP findings, statistically significant associations with plasma P-tau217 were observed for *APOE*-ε4 (β = 0.150, 95% CI: 0.098–0.202, ΔR² = 0.094), PGS_Amyloid-PET_ (β = 0.030, 95% CI: 0.004–0.056, ΔR² = 0.016), and PGS_CSF-P-tau181_ (β = 0.031, 95% CI: 0.006–0.057, ΔR² = 0.018), but no significant association was observed with PGS_Plasma-P-tau181_ and PGS_AD_ (Supplementary Figure 4). Age-stratified analyses indicated that associations with plasma P-tau217 were observed for *APOE*-ε4 and PGS_CSF-P-tau181_ both before and after age 70, whereas the association with PGS_Amyloid-PET_ and PGS_Plasma-P-tau181_ was only observed after age 70. No statistically significant interactions were observed between any of the genetic predictors and age group, sex, or *APOE*-ε4 carrier status. In addition to P-tau217, both *APOE*-ε4 and PGS_Amyloid-PET_ were associated with plasma Aβ42/Aβ40 and P-tau217/Aβ42; PGS_CSF-P-tau181_ and PGS_AD_ were only associated with P-tau217/Aβ42 (Supplementary Table 2). Second, we replicated the predictive value of Core-1 biomarkers for cognition in non-demented non-Hispanic white WHICAP participants younger than 85 years (N = 721, n = 2,386). PGS_Amyloid-PET_ was associated with global cognition and processing speed, whereas PGS_CSF-P-tau181_ was only associated with visuospatial ability, and PGS_Plasma-P-tau181_ was associated with language ability (Supplementary Table 3). Third, the AD PGS derived from the GWAS by Kunkle et al. showed limited predictive value for P-tau217 (results not shown, upon request), and the associations of individual genetic predictors with P-tau181 and P-tau217 remained consistent in the smaller WRAP cohort based on the complete case analysis (Supplementary Table 4). Among the EFIGA tuning cohort of Hispanic individuals (n = 305), the PGS_Plasma-P-tau181_ showed associations with plasma GFAP in the full sample, but not in the unaffected subgroup. When applied in the EFIGA tuning cohort, PGS_GAUDI_ was associated with plasma P-tau181 in both the full and unaffected cohorts and was marginally associated with plasma GFAP in the unaffected group (Supplementary Table 5).

### 3.7 Integrative Core-1 biomarker PGS

In WRAP (Table 5), PGS_Integrative_ showed significant associations with baseline (β = 0.030, 95% CI: 0.014–0.045, ΔR² = 0.017) and longitudinal P-tau217 levels (β = 0.030, 95% CI: 0.014–0.046, ΔR² = 0.013). For other biomarkers, significant associations were observed with P-tau217/Aβ42. For longitudinal cognition, associations were observed with executive function and PACC-3. Similar findings were observed in Wisconsin ADRC and WHICAP (Supplementary Table 6), where PGS_Integrative_ showed significant associations with P-tau217 (β = 0.046, 95% CI: 0.020–0.072, ΔR² = 0.039), P-tau217/Aβ42, and longitudinal cognition. In age-stratified analyses, PGS_Integrative_ showed at least marginal associations with P-tau217 across all age groups in both WRAP and the Wisconsin ADRC. Associations stratified by sex, and *APOE* allele status are shown in Supplementary Figure 5.

**Table 5.** Associations Of Integrative Core-1 Biomarker PGS With Plasma Biomarkers And Cognition Among Non-Hispanic White Individuals (n_max_ = 871*, N_max_ = 2,750) Table 5 presents the associations of the integrative Core-1 biomarker PGS with baseline and longitudinal P-tau217, other biomarkers, and cognition among non-Hispanic White individuals using the full WRAP cohort. The integrative Core-1 biomarker PGS was an unweighted composite of three individual Core-1 PGSs, which included PGSs for amyloid, CSF P-tau181, and plasma P-tau181. All individual PGSs were standardized before inclusion in the composite score. WRAP participants were restricted to those aged 62 and older. For baseline analyses, we used linear models. For all longitudinal analyses, we used linear mixed-effects models adjusted for age, sex, practice effects (only for cognition, visit number minus one), and the first 10 principal components, and included random intercepts for subjects to account for within-individual correlations. We excluded WRAP participants who were also included in the GWAS of amyloid PET to avoid sample overlap. The final integrative PGS was z-standardized. Bolded values indicate p < 0.05. n_max_: Maximum number of unique individuals included in the sample after excluding those who were also part of the discovery GWAS. The actual sample size varies by outcome, as some cognitive assessments and biomarkers were not administered to all participants. N_max_: Maximum number of person-level observations (i.e., person-years). The number of observations varies by outcome due to missing data at one or more follow-up visits. *In the full WRAP sample, 89 individuals overlapped between the amyloid GWAS and the target sample. After restricting the analysis to participants aged 62 or older, 84 of these individuals remained and were therefore excluded from downstream analyses.

| P-tau217 |  |  |  |  | Other biomarkers (longitudinal) |  |  |  | Cognition (longitudinal) |  |  |  |
| --- | --- | --- | --- | --- | --- | --- | --- | --- | --- | --- | --- | --- |
| Outcome | Beta | SE | P | $\Delta R^2$ | Outcome | Beta | SE | P | Outcome | Beta | SE | P |
| P-tau217 (baseline) | 0.030 | 0.008 | **<0.001** | 0.017 | Aβ42/Aβ40 | -0.006 | 0.004 | 0.152 | Delayed recall | -0.054 | 0.029 | 0.060 |
| P-tau217 (longitudinal) | 0.030 | 0.008 | **<0.001** | 0.013 | GFAP | 0.004 | 0.006 | 0.493 | Immediate learning | -0.042 | 0.028 | 0.139 |
|  |  |  |  |  | NfL | 0.000 | 0.006 | 0.946 | Executive function | -0.083 | 0.027 | **0.002** |
| | | | | | P-tau217/Aβ42 | 0.032 | 0.010 | **0.001** | PACC-3 | -0.060 | 0.028 | **0.030** |

## 4. DISCUSSION

The recently revised NIA-AA criteria[1] introduced Core-1 and Core-2 biomarkers, with Core-1 capturing the earliest stage of AD that can be detected in vivo and used diagnostically. Here, we computed PGS for Core-1 biomarkers, including amyloid PET, plasma P-tau181, and CSF P-tau181, and evaluated their ability to predict plasma P-tau217 and P-tau181. We also compared their performance to that of *APOE* and a clinical AD PGS, which reflects eventual AD diagnosis, across multiple outcomes - including non-tau plasma biomarkers and cognitive change - in older non-Hispanic white and Caribbean Hispanic individuals. We found that *APOE* was the strongest predictor of plasma P-tau217 in non-Hispanic white individuals but showed weaker associations with P-tau181 in both groups. Although individual PRSs remained limited in predicting plasma P-tau181 and P-tau217, Core-1 biomarker PGSs generally outperformed the clinical AD PGS in predicting longitudinal changes in plasma biomarkers and cognition among asymptomatic individuals from both population sources. We also observed that an integrative Core-1 PGS may better predict P-tau217 levels among non-Hispanic white individuals.

The relationship between PGS and plasma P-tau181 or P-tau217 remains unclear, with few studies using AD GWAS-derived PGSs showing limited predictive value. In ADNI, a significant association between AD PGS and plasma P-tau181 was found, but only among individuals with mild cognitive impairment; no association was observed among cognitively intact individuals or those with AD[5]. A European cohort of similar size failed to detect any significant association between AD PGS and plasma P-tau181[6]. A second European study extended this analysis to plasma P-tau217 but also found no significant association with AD PGS[7], consistent with our results. This indicates that AD PGSs based on clinical case-control definitions may have limited utility in predicting biological AD. The reduced predictive power of these PGS for P-tau181 and P-tau217 is likely due to two factors. First, variants identified through clinical AD GWASs may reflect broader disease mechanisms rather than those directly related to early amyloid and tau pathology[8]. Supporting this, no association was found between an AD PGS and amyloid PET when using all AD-associated variants, but they observed a significant association when restricting the analysis to variants also linked to amyloid burden[9]. Second, reduced predictive power may result from misalignment between clinical AD diagnosis and biomarker-defined AD. Nearly 50% of individuals clinically diagnosed with AD can be biomarker negative, whereas approximately 20% of clinically non-demented individuals were biomarker positive[43].

*APOE*-ε4 remained a strong predictor of plasma P-tau217. Among non-Hispanic white individuals, *APOE-*ε4 alone explained 6–9% of the variance in plasma P-tau217 levels, consistent with findings from a recent study[7]. However, its predictive power was notably reduced for plasma P-tau181. *APOE*-ε4 accounted for only 1.4% to 2.4% of the variance in plasma P-tau181 among non-Hispanic white individuals, and less than 1% among Hispanic individuals, consistent with the weak association reported in a recent European study[6]. Recent GWASs of plasma P-tau181 using ADNI data have also been inconsistencies in the effect size of *APOE*[10,12]. Among non-Hispanic white individuals in our study, the lower predictive power of *APOE*-ε4 for P-tau181 compared to P-tau217 may reflect the greater sensitivity and specificity of P-tau217 for AD pathology[44]. The reduced predictive value for P-tau181 may also be partly explained by our focus on asymptomatic individuals in WRAP, as previous studies have shown weaker associations between *APOE* and P-tau181 in cognitively intact populations[5].

PGS derived from Core-1 biomarkers generally outperformed the clinical AD PGS in predicting P-tau181 and P-tau217 levels. However, the additional variance explained remained modest, and the degree of improvement appeared to vary by age. For PGS_Amyloid-PET_, associations were stronger among individuals older than 70 years and were observed consistently in both WRAP and Wisconsin-ADRC. In contrast, associations for PGS_CSF-P-tau181_, PGS_Plasma-P-tau181_, and PGS_AD_ appeared stronger among individuals younger than 70 years, with the association for PGS_Plasma-P-tau181_ in the younger group observed in both non-Hispanic white and Hispanic populations. However, these age-related associations for PGS_CSF-P-tau181_, PGS_Plasma-P-tau181_ and PGS_AD_ were not replicated. In Hispanic individuals, the PGS_GAUDI_ trained on P-tau181 outperformed traditional C+T methods in predicting P-tau181 levels, potentially reflecting ancestry-specific effects related to this biomarker[40]. However, this finding requires validation in an independent sample.

PGSs derived from Core-1 biomarkers showed stronger predictive value for preclinical changes in cognition and AD plasma biomarkers among asymptomatic individuals, particularly those based on amyloid PET and CSF P-tau181. For longitudinal cognition, PGSs derived from amyloid PET and CSF P-tau181 were at least marginally associated with a decline across multiple cognitive domains in WRAP. Although the PGS based on plasma P-tau181 did not significantly predict cognitive decline among non-Hispanic white individuals, it was associated with cognitive outcomes among Hispanic individuals. Similar findings were replicated in non-Hispanic white individuals from WHICAP. These results are consistent with previous findings from ADNI and Anti-Amyloid Treatment in Asymptomatic Alzheimer’s, where improved prediction of cognitive outcomes was also observed for Core-1 PGSs derived from CSF Aβ42 and P-tau181[45,46].

For other plasma biomarkers, we only observed consistent associations across WRAP and Wisconsin-ADRC for the PGS derived from amyloid PET, which was significantly associated with Aβ-related outcomes, including plasma Aβ42/Aβ40 and P-tau217/Aβ42—aligning with its underlying pathology. Among Hispanic individuals, consistent associations with GFAP were observed only for the PGS derived from plasma P-tau181, with replication in a subsample of EFIGA, although the strength of the association varied depending on the PGS construction method. These findings are in line with a recent study suggesting that plasma GFAP is an early marker of brain Aβ pathology, rather than tau aggregation[47].

Despite the modest performance of individual Core-1 biomarker PGS among non-Hispanic white individuals, the low correlations among the top-performing PGSs derived from amyloid PET, plasma P-tau181, and CSF P-tau181 suggest that combining these scores may improve predictive power. The integrative PGS showed improved predictive performance for plasma P-tau217 and reduced the age-related differences observed with individual PRSs. This improvement aligns with recent literature on cross-trait PGS prediction, which suggests that combining PGSs for multiple disease-related risk factors or subtypes can offer greater predictive value than using individual PGSs alone[48].

In the present analysis, we constructed an integrative Core-1 PGS using only three Core-1 biomarkers in an unweighted composite yet observed improved performance. However, according to existing guidelines, there are over a dozen of Core-1 biomarkers—measurable in plasma, CSF, or via PET (including relevant ratios)—that could potentially be incorporated. Moreover, applying advanced methods to combine these biomarkers—such as linear combinations or elastic net regression—may further enhance predictive accuracy. An integrative PGS combining diverse biomarkers, clinical phenotypes, and advanced methods may improve prediction of both clinically defined and biomarker-defined AD. However, such integrative PGSs may not be predictive for a specific biomarker or endophenotype. In contrast, integrative PGSs optimized for specific outcomes (e.g., cognition or individual biomarkers) by including PGSs of relevant endophenotypes may offer enhanced predictive power for those targeted phenotypes.

Our study has limitations. First, the GWAS for Core-1 biomarkers used in the current analyses were relatively small, partly due to difficulties in accessing summary statistics from larger published studies. Small GWAS may introduce greater uncertainty in PGS estimates and lead to unstable performance due to statistical noise. Although we partially addressed this through internal replication, the replication datasets were modest in size and may have lacked sufficient power to replicate certain associations with AD endophenotypes. Moreover, the internal replications were not fully independent, as some datasets (e.g., EFIGA and Wisconsin-ADRC) were also used for PGS tuning. Taken together, the findings should be interpreted with caution due to overfitting and potential inflation of effect estimates. Second, although we derived PGS for amyloid PET, CSF P-tau181, and plasma P-tau181, we were unable to include a PGS based on plasma P-tau217 due to the lack of available data for both GWAS training and PGS testing. A PGS derived directly from plasma P-tau217 may have stronger predictive value for this biomarker compared to PGSs based on CSF or plasma P-tau181. Third, we constructed PGS primarily using traditional C+T methods; other PGS approaches may further improve predictive performance. Fourth, we did not apply multiple testing correction due to concerns about missing potentially true signals, as most observed effects were weak. Instead, we focused on replicating key findings in independent datasets to support their robustness. Fifth, we tuned the PGS parameters using plasma P-tau217/P-tau181 as the outcome. However, the results may be inconsistent when different biomarkers are used for tuning (e.g., plasma Aβ42/Aβ40). Sixth, longitudinal data availability varied across outcomes, and a global complete case analysis was not conducted due to power concerns, potentially limiting result comparability. Seventh, the small size of the study of P-tau181 among non-Hispanic white individuals may reduce the generalizability of the findings.

Together, our findings suggest that individual and integrative PGSs derived from Core-1 biomarkers may have utility in predicting plasma P-tau181 and P-tau217, as well as other biomarkers and cognitive outcomes, among asymptomatic individuals across populations. These findings support the identification of at-risk individuals for early biomarker screening and clinical trial recruitment, contributing to ongoing efforts to enhance trial efficacy and improve disease stratification for biologically defined AD.

## Data availability statement

The data analyzed in this study are subject to the following licenses/restrictions: datasets can be requested through formal research applications to the Washington Heights-Inwood Columbia Aging Project and the Genetic Studies of Alzheimer’s Disease in Caribbean Hispanics (EFIGA). Requests to access these datasets should be directed to the Columbia University Alzheimer’s Disease Research Center: https://www.neurology.columbia.edu/research/research-centers-and-programs/alzheimers-disease-research-center-adrc/investigators/investigator-resources.

Data may also be requested from the Wisconsin Registry for Alzheimer’s Prevention and the Wisconsin Alzheimer’s Disease Research Center via: https://wrap.wisc.edu/data-requests-2/, and from the Alzheimer’s Disease Neuroimaging Initiative (ADNI): https://adni.loni.usc.edu/data-samples/adni-data/.

## Ethics statement

The studies involving humans were approved by the Institutional Review Board of the Columbia University Medical Center. The studies were conducted in accordance with the local legislation and institutional requirements. Subjects from WRAP, Wisconsin-ADRC, WHICAP, and EFIGA have provided signed informed consent before participation.

## Author contributions

YX and RM contributed to the study design. YX, RM, DR, LH, and YG prepared the data and performed the data analysis. YX and RM drafted the manuscript. All authors critically reviewed the manuscript and have approved the final version.

## Acknowledgements

We acknowledge the participants from the Washington Heights-Inwood Columbia Aging Project (WHICAP), the Genetic Studies of Alzheimer’s Disease in Caribbean Hispanics (EFIGA), the Wisconsin Registry for Alzheimer’s Prevention (WRAP), the Wisconsin Alzheimer’s Disease Research Center (Wisconsin ADRC), and the Alzheimer’s Disease Neuroimaging Initiative (ADNI) for their invaluable contributions and dedication. This study would not be possible without their continued participation. We also thank the researchers and study teams involved in WHICAP, EFIGA, WRAP, the Wisconsin ADRC, and ADNI for their longstanding efforts in data collection and subject follow-up over the years.

