## Supplementary materials for "Polygenic Scores of Core-1 Alzheimer’s Disease Biomarkers Predict Early Cognitive and Pathological Change"

**Correspondence:**

Richard P. Mayeux, MD

**Supplementary methods**

*Sample description*

Wisconsin Registry for Alzheimer’s Prevention: The Wisconsin Registry for Alzheimer’s Prevention (WRAP) is an ongoing, prospective, longitudinal cohort study of over 1,500 cognitively healthy, English-speaking adults who were between 40 and 65 years old at baseline [1]. The WRAP cohort is intentionally enriched for individuals with a parental history of Alzheimer’s disease (AD), thereby increasing the likelihood of observing late-onset AD pathology and cognitive decline over time. More than 70% of participants had a parent with autopsy-confirmed or probable AD, as defined by the National Institute of Neurological and Communicative Disorders and Stroke and the Alzheimer's Disease and Related Disorders Association (NINCDS-ADRDA) criteria. Recruitment began in 2001, with an initial follow-up after four years and subsequent visits conducted every two years.

Wisconsin Alzheimer’s Disease Research Center: The Wisconsin Alzheimer’s Disease Research Center (Wisconsin-ADRC) was established in 2009 and is one of the National Institute on Aging (NIA)-designated ADRCs across the United States [2]. The Wisconsin ADRC follows participants longitudinally. Adults aged 45 and older with decisional capacity and English fluency were eligible for enrollment. Exclusion criteria included active major medical or psychiatric illness and the absence of a study partner.

Alzheimer’s Disease Neuroimaging Initiative: The Alzheimer’s Disease Neuroimaging Initiative (ADNI), launched in 2003 as a public–private partnership under the leadership of Principal Investigator Michael W. Weiner, MD, aims to determine whether serial magnetic resonance imaging (MRI), PET imaging, other biological markers, and clinical and neuropsychological assessments can be integrated to track the progression of mild cognitive impairment (MCI) and early Alzheimer’s disease (AD) [3]. For the most current information, visit [www.adni-info.org](http://www.adni-info.org).

Washington Heights-Hamilton Heights-Inwood Columbia Aging Project (WHICAP): The Washington Heights-Inwood Columbia Aging Project (WHICAP) is a multi-ethnic, community-based, prospective cohort study investigating clinical and genetic risk factors for dementia [4]. Participants were enrolled in three waves (1992, 1999, and 2009), all following consistent study protocols. Individuals were recruited to represent older adults residing in the northern Manhattan community. At study entry, participants completed a structured interview assessing general health and functional status, followed by a comprehensive evaluation that included medical, neurological, and psychiatric histories, as well as standardized physical, neurological, and neuropsychological examinations. Follow-up visits were conducted every 18 to 24 months using procedures similar to those at baseline.

Estudio Familiar de Influencia Genética en Alzheimer: The Estudio Familiar de Influencia Genética en Alzheimer (EFIGA) began in 1998 and focuses on individuals of Caribbean Hispanic descent, including those with familial and sporadic late-onset AD, as well as cognitively healthy, non-demented individuals from the Dominican Republic and New York [5]. EFIGA collects detailed information on dementia status, general health, and functional abilities, adhering to the same diagnostic criteria and protocols as WHICAP.

*Biomarker acquisition and processing*

Plasma biomarkers for WRAP and the Wisconsin ADRC were analyzed at the University of Gothenburg [6]. The primary biomarker of interest, P-tau217, was measured in duplicate using a commercially available ALZpath assay (ALZpathDX, Carlsbad, CA), known for its high sensitivity, precision, and reproducibility. Additional biomarkers—including Aβ42/40, P-tau181, GFAP, and NfL—were quantified from the same aliquots using Quanterix kits and in-house Simoa assays. All biomarkers were z-standardized relative to the AD-negative cognitively unimpaired control group.

CSF samples from ADNI, collected according to the procedures outlined in the ADNI procedures manual (http://www.adni-info.org), were analyzed using Elecsys CSF immunoassays on a cobas e 601 analyzer at the University of Pennsylvania [7,8]. Plasma P-tau181 was measured using the single molecule array (Simoa) technique with an in-house assay developed by the Clinical Neurochemistry Laboratory at the University of Gothenburg [9,10]. CSF and plasma data were downloaded from the ADNI database (https://ida.loni.usc.edu) in early 2025.

Blood samples from WHICAP and EFIGA participants were collected via standard venipuncture into dipotassium EDTA tubes [4]. Plasma was isolated by centrifugation at 2,000 × g for 15 minutes at 4°C within two hours of collection, aliquoted into polypropylene tubes, and stored at −80°C. Plasma biomarker assays were conducted between April and November 2022 using Quanterix Simoa (single molecule array) technology on the HD-X platform (Quanterix, Billerica, MA, USA). Samples were diluted and analyzed in duplicate according to the manufacturer’s instructions, utilizing three Quanterix kits: Neurology 3-Plex A (Cat. No. 101995) for Aβ42, Aβ40, and total tau; P-tau181 V2 Advantage (Cat. No. 103714) for tau phosphorylated at threonine 181 (p-tau181); and Neurology 2-Plex B (Cat. No. 103520) for NfL and GFAP.

**Supplementary Table 1. Descriptive Statistics For The Samples Used In GWAS And PGS Tuning**

|  | | **NHW** | | **CH** | |
| --- | --- | --- | --- | --- | --- |
| **Characteristic** | **GWAS:**  **ADNI CSF** n = 830*^1^* | **GWAS:**  **ADNI Plasma** n = 793*^1^* | **PGS Tuning:**  **Wisconsin-ADRC** n = 301*^1^* | **GWAS:**  **EFIGA** n = 1,220*^1^* | **PGS Tuning:**  **EFIGA** n = 305*^1^* |
| Age (years) | 75.11 (7.50) | 77.73 (7.65) | 69.12 (8.44) | 72.14 (8.20) | 72.22 (8.19) |
| ***Sex*** |  |  |  |  |  |
| Men | 465 (56%) | 441 (56%) | 115 (38%) | 393 (32%) | 107 (35%) |
| Women | 365 (44%) | 352 (44%) | 186 (62%) | 827 (68%) | 198 (65%) |
| Education (years) | 16.03 (2.83) | 16.11 (2.78) | 16.78 (2.31) | 5.76 (5.37) | 6.12 (5.37) |
| AD | 120 (14%) | 47 (6%) | 54 (18%) | 321 (26%) | 70 (23%) |
| ***APOE*** |  |  |  |  |  |
| 2\2 | 1 (0%) | 3 (0%) | 2 (1%) | 5 (0%) | 1 (0%) |
| 2\3 | 70 (8%) | 71 (9%) | 36 (12%) | 131 (11%) | 39 (13%) |
| 2\4 | 10 (1%) | 15 (2%) | 11 (4%) | 32 (3%) | 3 (1%) |
| 3\3 | 400 (48%) | 406 (51%) | 137 (46%) | 672 (55%) | 153 (50%) |
| 3\4 | 270 (33%) | 240 (30%) | 93 (31%) | 328 (27%) | 91 (30%) |
| 4\4 | 79 (10%) | 58 (7%) | 22 (7%) | 47 (4%) | 16 (5%) |
| *^1^* Mean (SD); n (%) | | | | | |

NHW: Non-Hispanic White; CH: Caribbean Hispanics.

**Supplementary Table 2. Associations of Genetic Predictors with Longitudinal Plasma Biomarkers Among Non-Hispanic Whites in Wisconsin-ADRC (n_max_ = 247, N_max_ = 299)**

| **Predictor** | **Aβ42/Aβ40** | **GFAP** | **NfL** | **P-tau217/Aβ42** |
| --- | --- | --- | --- | --- |
| *APOE* ε4 | **-0.043 (-0.080–-0.005) p = 0.03** | 0.020 (-0.039–0.078) p = 0.51 | -0.005 (-0.057–0.048) p = 0.86 | **0.220 (0.158–0.281) p < 0.01** |
| Amyloid PGS (C+T) | **-0.022 (-0.039–-0.004) p = 0.02** | -0.007 (-0.035–0.020) p = 0.59 | -0.005 (-0.029–0.020) p = 0.71 | **0.045 (0.014–0.076) p < 0.01** |
| CSF P-tau181 PGS (C+T) | -0.007 (-0.025–0.010) p = 0.41 | -0.015 (-0.042–0.012) p = 0.29 | -0.012 (-0.036–0.012) p = 0.33 | **0.036 (0.005–0.067) p = 0.02** |
| Plasma P-tau181 PGS (C+T) | 0.014 (-0.003–0.031) p = 0.10 | 0.005 (-0.021–0.031) p = 0.70 | -0.015 (-0.039–0.008) p = 0.20 | 0.012 (-0.019–0.042) p = 0.44 |
| AD PGS (C+T) | 0.003 (-0.015–0.022) p = 0.72 | -0.011 (-0.040–0.018) p = 0.44 | -0.015 (-0.041–0.010) p = 0.24 | 0.032 (-0.001–0.065) p = 0.05 |

**Supplementary Table 2** presents the longitudinal associations of *APOE* ε4, Core-1 biomarker-derived PGSs, and the AD GWAS PGS with plasma biomarkers beyond P-tau217 among non-demented, non-Hispanic White individuals in the Wisconsin ADRC. All longitudinal analyses were conducted using linear mixed-effects models adjusted for age, sex, and the first 10 principal components. Random intercepts were included to account for within-individual correlations. All continuous genetic predictors were z-standardized. Ninety-five percent confidence intervals are shown in parentheses, and bolded values indicate p < 0.05. C+T: clumping and thresholding; n_max_: Maximum number of unique individuals included in the sample. Actual sample size varies by outcome. N_max_: Maximum number of person-level observations (i.e., person-years). The number of observations varies by outcome due to missing biomarker data at one or more follow-up visits.

**Supplementary Table 3. Associations of Genetic Predictors with Longitudinal Cognition Among Non-Hispanic Whites in WHICAP (n_max_ = 721, N_max_ = 2,386)**

| **Predictor** | **Global** | **Memory** | **Speed** | **Visuospatial** | **Language** |
| --- | --- | --- | --- | --- | --- |
| *APOE* ε4 | -0.010 (-0.063–0.043) p = 0.72 | 0.011 (-0.070–0.093) p = 0.78 | -0.062 (-0.160–0.036) p = 0.21 | -0.008 (-0.054–0.037) p = 0.72 | 0.045 (-0.024–0.114) p = 0.20 |
| Amyloid PGS (C+T) | **-0.025 (-0.048–-0.003) p = 0.03** | -0.012 (-0.047–0.023) p = 0.50 | **-0.042 (-0.085–-0.000) p = 0.05** | -0.002 (-0.021–0.018) p = 0.86 | -0.012 (-0.041–0.018) p = 0.43 |
| CSF P-tau181 PGS (C+T) | -0.033 (-0.078–0.012) p = 0.15 | -0.038 (-0.105–0.029) p = 0.27 | -0.056 (-0.140–0.027) p = 0.19 | **-0.044 (-0.082–-0.007) p = 0.02** | -0.035 (-0.092–0.022) p = 0.22 |
| Plasma P-tau181 PGS (C+T) | -0.008 (-0.032–0.015) p = 0.48 | -0.006 (-0.041–0.029) p = 0.75 | -0.036 (-0.079–0.007) p = 0.10 | 0.001 (-0.018–0.021) p = 0.89 | **-0.030 (-0.060–-0.000) p = 0.05** |

**Supplementary Table 3** presents the longitudinal associations of *APOE* ε4, Core-1 biomarker-derived PGSs, and the AD GWAS PGS with cognitive outcomes among non-demented, non-Hispanic White individuals in WHICAP. Because WHICAP participants are older than those in the main analysis, we restricted the WHICAP sample to non-demented individuals younger than age 85 to improve comparability. PGS_AD_ was not evaluated in WHICAP because the WHICAP sample was included in the AD GWAS meta-analysis, and we were unable to identify the specific individuals from WHICAP who were part of that GWAS. All longitudinal analyses were conducted using linear mixed-effects models adjusted for age, sex, practice effects (visit number minus one), and the first 10 principal components. Random intercepts were included to account for within-individual correlations. All continuous genetic predictors were z-standardized. Ninety-five percent confidence intervals are shown in parentheses, and bolded values indicate p < 0.05. C+T: clumping and thresholding; n_max_: Maximum number of unique individuals included in the sample. Actual sample size varies by outcome. N_max_: Maximum number of person-level observations (i.e., person-years). The number of observations varies by outcome due to missing cognition data at one or more follow-up visits.

**Supplementary Table 4. Associations Of Individual Genetic Predictors With Plasma P-Tau181 And P-Tau217 In The Smaller WRAP Sample (Complete-Case Analysis) (n_max_ = 325, N_max_ = 798)**

| **P-tau217** | | | | **P-tau181** | | | |
| --- | --- | --- | --- | --- | --- | --- | --- |
| **Outcome** | **Beta** | **SE** | **P** | **Outcome** | **Beta** | **SE** | **P** |
| **Baseline** |  |  |  |  |  |  |  |
| *APOE* ε4 | 0.140 | 0.026 | **<0.001** | *APOE* ε4 | 0.061 | 0.023 | **0.007** |
| Amyloid PGS | 0.027 | 0.022 | 0.230 | Amyloid PGS | 0.047 | 0.020 | **0.019** |
| CSF P-tau181 PGS | 0.009 | 0.013 | 0.484 | CSF P-tau181 PGS | 0.020 | 0.011 | 0.080 |
| Plasma P-tau181 PGS | 0.016 | 0.014 | 0.232 | Plasma P-tau181 PGS | 0.018 | 0.011 | 0.124 |
| AD PGS | 0.011 | 0.013 | 0.415 | AD PGS | 0.006 | 0.011 | 0.604 |
| **Longitudinal** |  |  |  |  |  |  |  |
| *APOE* ε4 | 0.169 | 0.026 | **<0.001** | *APOE* ε4 | 0.077 | 0.021 | **<0.001** |
| Amyloid PGS | 0.032 | 0.020 | 0.110 | Amyloid PGS | 0.029 | 0.016 | 0.073 |
| CSF P-tau181 PGS | 0.016 | 0.013 | 0.209 | CSF P-tau181 PGS | 0.010 | 0.011 | 0.376 |
| Plasma P-tau181 PGS | 0.012 | 0.013 | 0.346 | Plasma P-tau181 PGS | 0.020 | 0.011 | 0.067 |
| AD PGS | 0.006 | 0.013 | 0.648 | AD PGS | 0.004 | 0.011 | 0.733 |

**Supplementary Table 4** presents the association of individual genetic predictors with plasma P-tau181 and P-tau217 in the WRAP smaller sample with complete-case analysis. All PGS predictors were first tuned in the Wisconsin ADRC cohort, and the best-performing PGSs for P-tau217 were then applied to WRAP. To ensure comparability with the Caribbean Hispanic sample, we restricted the WRAP sample to participants aged ≥62 and used the first visit after age 62 as the baseline. Baseline association was estimated using linear regression adjusting for age, sex, and the first 10 PCs, and longitudinal association was estimated using linear mixed models adjusting for the same covariates, with random intercepts. For PGSs derived from the amyloid PET GWAS, we excluded 89 WRAP participants who overlapped with the discovery GWAS. All continuous predictors were z-standardized; n_max_: Maximum number of unique individuals included in the sample. Actual sample size varies by predictor due to exclusion of individuals who were also part of the discovery GWAS. N_max_: Maximum number of person-level observations (i.e., person-years).

**Supplementary Table 5. Associations of Genetic Predictors with Longitudinal Plasma Biomarkers Among Caribbean Hispanics in a subset of EFIGA (n_max_ = 305)**

|  | **P-tau181** | **Aβ42/Aβ40** | **GFAP** | **NfL** | **P-tau181/Aβ42** |
| --- | --- | --- | --- | --- | --- |
| **Full cohort (N = 305)** | | | | | |
| *APOE* ε4 | 0.048 (-0.002–0.098) p = 0.06 | -0.031 (-0.066–0.004) p = 0.08 | **0.081 (0.025–0.137) p < 0.01** | 0.062 (-0.008–0.131) p = 0.08 | **0.070 (0.009–0.131) p = 0.02** |
| Plasma P-tau181 PGS (GAUDI) | **0.029 (0.008–0.051) p < 0.01** | 0.004 (-0.012–0.021) p = 0.60 | 0.017 (-0.010–0.043) p = 0.22 | -0.013 (-0.042–0.016) p = 0.39 | 0.012 (-0.017–0.040) p = 0.41 |
| Plasma P-tau181 PGS (C+T) | 0.012 (-0.013–0.037) p = 0.34 | 0.016 (-0.004–0.035) p = 0.11 | **0.031 (0.000–0.062) p = 0.05** | 0.018 (-0.015–0.052) p = 0.28 | -0.009 (-0.042–0.024) p = 0.58 |
| AD PGS (C+T) | -0.002 (-0.030–0.026) p = 0.90 | 0.008 (-0.014–0.031) p = 0.46 | 0.007 (-0.029–0.044) p = 0.69 | 0.034 (-0.005–0.073) p = 0.08 | -0.018 (-0.055–0.020) p = 0.36 |
| **Unaffected (N = 235)** | | | | | |
| *APOE* ε4 | 0.009 (-0.047–0.066) p = 0.75 | -0.038 (-0.082–0.007) p = 0.10 | 0.051 (-0.016–0.118) p = 0.13 | 0.054 (-0.029–0.137) p = 0.20 | 0.031 (-0.041–0.103) p = 0.40 |
| Plasma P-tau181 PGS (GAUDI) | **0.031 (0.006–0.056) p = 0.01** | 0.013 (-0.008–0.035) p = 0.22 | 0.025 (-0.003–0.053) p = 0.08 | -0.030 (-0.070–0.009) p = 0.13 | 0.007 (-0.027–0.042) p = 0.68 |
| Plasma P-tau181 PGS (C+T) | 0.020 (-0.009–0.048) p = 0.17 | 0.017 (-0.005–0.039) p = 0.14 | 0.015 (-0.014–0.044) p = 0.32 | 0.025 (-0.017–0.066) p = 0.24 | -0.003 (-0.039–0.033) p = 0.87 |
| AD PGS (C+T) | -0.013 (-0.048–0.021) p = 0.45 | 0.009 (-0.019–0.038) p = 0.51 | -0.019 (-0.056–0.019) p = 0.32 | 0.042 (-0.011–0.096) p = 0.12 | -0.029 (-0.074–0.016) p = 0.20 |

**Supplementary Table 5** presents the cross-sectional associations of *APOE* ε4, Core-1 biomarker-derived PGSs, and the AD GWAS PGS with plasma biomarkers in the EFIGA tuning sample. Due to the cross-sectional nature of the data and the small sample size, we present results for both the full sample and the subset of non-demented individuals. All analyses were conducted using linear models adjusted for age, sex, and the first 10 principal components. For PGS_AD_ analyses, we excluded 30 participants who were also included in the AD GWAS. All continuous genetic predictors were z-standardized. Ninety-five percent confidence intervals are shown in parentheses, and bolded values indicate p < 0.05. C+T: clumping and thresholding; GAUDI: a powerful tool for PGS analysis in admixed populations, as it explicitly models ancestry-differential effects while borrowing information across segments with shared ancestry; n_max_: Maximum number of unique individuals included in the sample. Actual sample size varies by outcome/predictor due to exclusion of individuals who were also part of the discovery GWAS.

**Supplementary Table 6. Associations Of Integrative Core-1 Biomarker PGS With Plasma Biomarkers And Cognition Among Non-Hispanic White Individuals (From WHICAP And Wisconsin ADRC)**

| **Longitudinal cognition (WHICAP)**  **(n_max_ = 721, N_max_ = 2,386)** | | | | **Longitudinal biomarker (Wisconsin-ADRC)**  **(n_max_ = 247, N_max_ = 299)** | | | |
| --- | --- | --- | --- | --- | --- | --- | --- |
| **Outcome** | **Beta** | **SE** | **P** | **Outcome** | **Beta** | **SE** | **P** |
| Memory | -0.018 | 0.019 | 0.325 | P-tau217 | 0.046 | 0.013 | **0.001** |
| Speed | -0.062 | 0.023 | **0.007** | Aβ42/Aβ40 | -0.007 | 0.009 | 0.424 |
| Visuospatial | -0.008 | 0.010 | 0.432 | GFAP | -0.009 | 0.014 | 0.525 |
| Language | -0.034 | 0.016 | **0.033** | NfL | -0.018 | 0.013 | 0.158 |
| Global | -0.029 | 0.012 | **0.021** | P-tau217/Aβ42 | 0.051 | 0.016 | **0.002** |

**Supplementary Table 6** presents the associations of the integrative Core-1 biomarker PGS with longitudinal biomarkers and cognition among non-Hispanic White individuals, using data from the WHICAP and Wisconsin ADRC cohorts. The integrative Core-1 biomarker PGS was an unweighted composite of three individual Core-1 PGSs, including PGSs for amyloid, CSF P-tau181, and plasma P-tau181. All individual PGSs were standardized before inclusion in the composite score. For all longitudinal analyses, we used linear mixed-effects models adjusted for age, sex, practice effects (for cognition only, defined as visit number minus one), and the first 10 principal components. Random intercepts for subjects were included to account for within-individual correlations. The final integrative PGS was z-standardized. Bolded values indicate p < 0.05; n_max_: Maximum number of unique individuals included in the sample. Actual sample size varies by outcome. N_max_: Maximum number of person-level observations (i.e., person-years). The number of observations varies by outcome due to missing biomarker/cognition data at one or more follow-up visits.

**Supplementary Figure 1. Correlations Among All Genetic Predictors In Non-Hispanic White And Caribbean Hispanic Individuals**

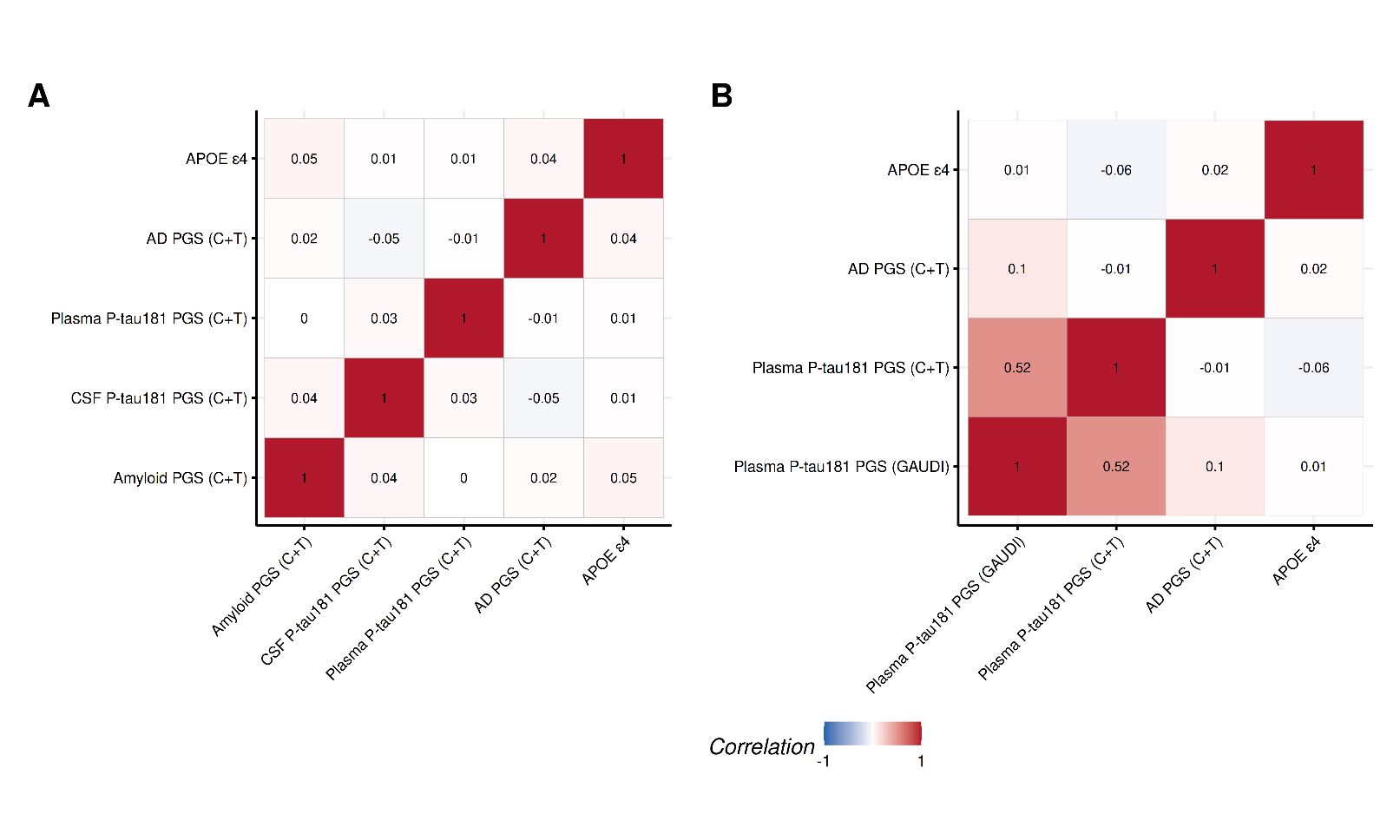

**Supplementary Figure 1** presents the pair-wise correlations among *APOE* ε4, Core-1 biomarker-derived PGSs, and the AD GWAS PGS. Correlation analyses for non-Hispanic White individuals were conducted in WRAP (Panel A), and analyses for Caribbean Hispanic individuals were conducted in WHICAP (Panel B). Blue indicates negative correlations, and red indicates positive correlations. The correlation coefficient is shown within each cell. C+T: clumping and thresholding; GAUDI: a powerful tool for PGS analysis in admixed populations that explicitly models ancestry-differential effects while borrowing information across segments with shared ancestry.

**Supplementary Figure 2. Age-Stratified Longitudinal Associations of *APOE* ε4 and PGSs Derived from GWAS of Core-1 Biomarkers and Clinical AD with Plasma P-tau181 and P-tau217 Among Non-Hispanic White and Hispanic Individuals (***See main text for sample-specific sizes)*

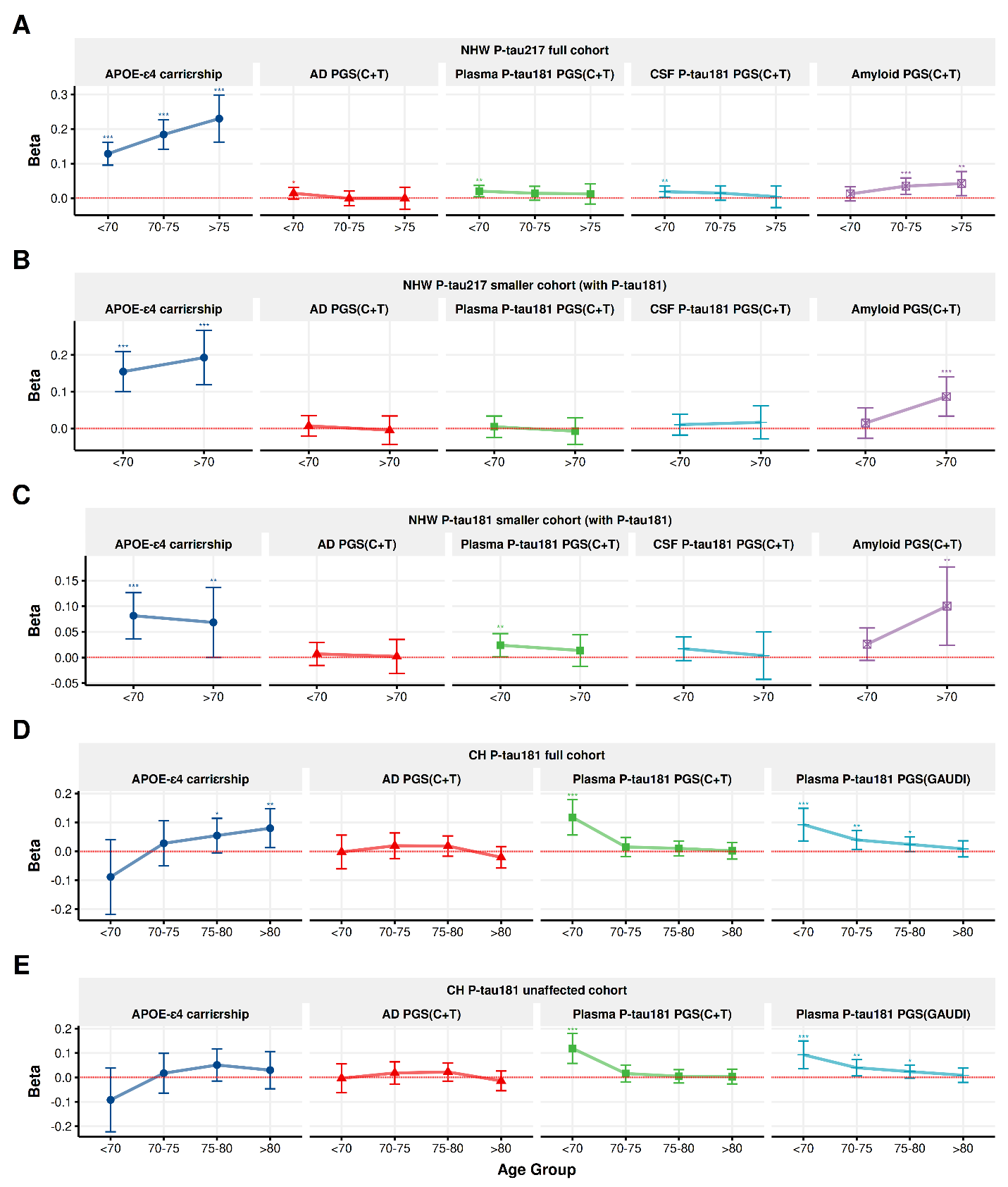

**Supplementary Figure 2** presents the age-stratified longitudinal associations of *APOE* ε4 and PGSs derived from GWAS of Core-1 biomarkers and clinical AD with plasma P-tau181 and P-tau217 among non-Hispanic white and Hispanic individuals. For the age group analyses, participants in the full WRAP sample were categorized as <70, 70–75, or >75 years old, while those in the smaller sample were grouped as <70 or >70 years old due to limited follow-up data at older ages. In WHICAP, we applied the same age stratification scheme as WRAP, with the addition of an >80 group to reflect the relatively older age distribution of the cohort. Panel A shows the association and predictive performance of all genetic predictors with plasma P-tau217 in the full WRAP sample. Panel B presents the same metrics with plasma P-tau217 in the WRAP subsample where P-tau181 is available. Panel C shows the association and predictive performance with plasma P-tau181 in WRAP. Panel D shows the association and predictive performance with plasma P-tau181 among Caribbean Hispanics in the full WHICAP sample, and Panel E presents the same metrics among non-demented Caribbean Hispanic individuals in WHICAP. All PGS predictors were first tuned in a designated tuning sample, and the PGSs that showed the best prediction performance in that sample were applied to the target samples. For the longitudinal analyses, we used linear mixed-effects models adjusting for age, sex, and the first 10 principal components, and included random intercepts for individuals to account for within-subject correlations. For analyses involving the PGS derived from amyloid PET imaging, we additionally excluded 89 WRAP participants who were included in the original GWAS to avoid sample overlap. For analyses involving the PGS derived from AD GWAS in the Hispanic sample, we excluded 219 WHICAP participants who were also included in the original GWAS to avoid sample overlap. All continuous genetic predictors were z-standardized. ***p < 0.01, **p < 0.05, *p < 0.1. C+T: clumping and thresholding; GAUDI: a powerful tool for PGS analysis in admixed populations, as it explicitly models ancestry-differential effects while borrowing information across segments with shared ancestry; NHW: Non-Hispanic Whites; CH: Caribbean Hispanics.

**Supplementary Figure 3. Sex- And *APOE* ε4-Stratified Longitudinal Associations Of Different Genetic Predictors With Plasma P-Tau181 And P-Tau217 Among White And Hispanic Individuals (***See main text for sample-specific sizes)*

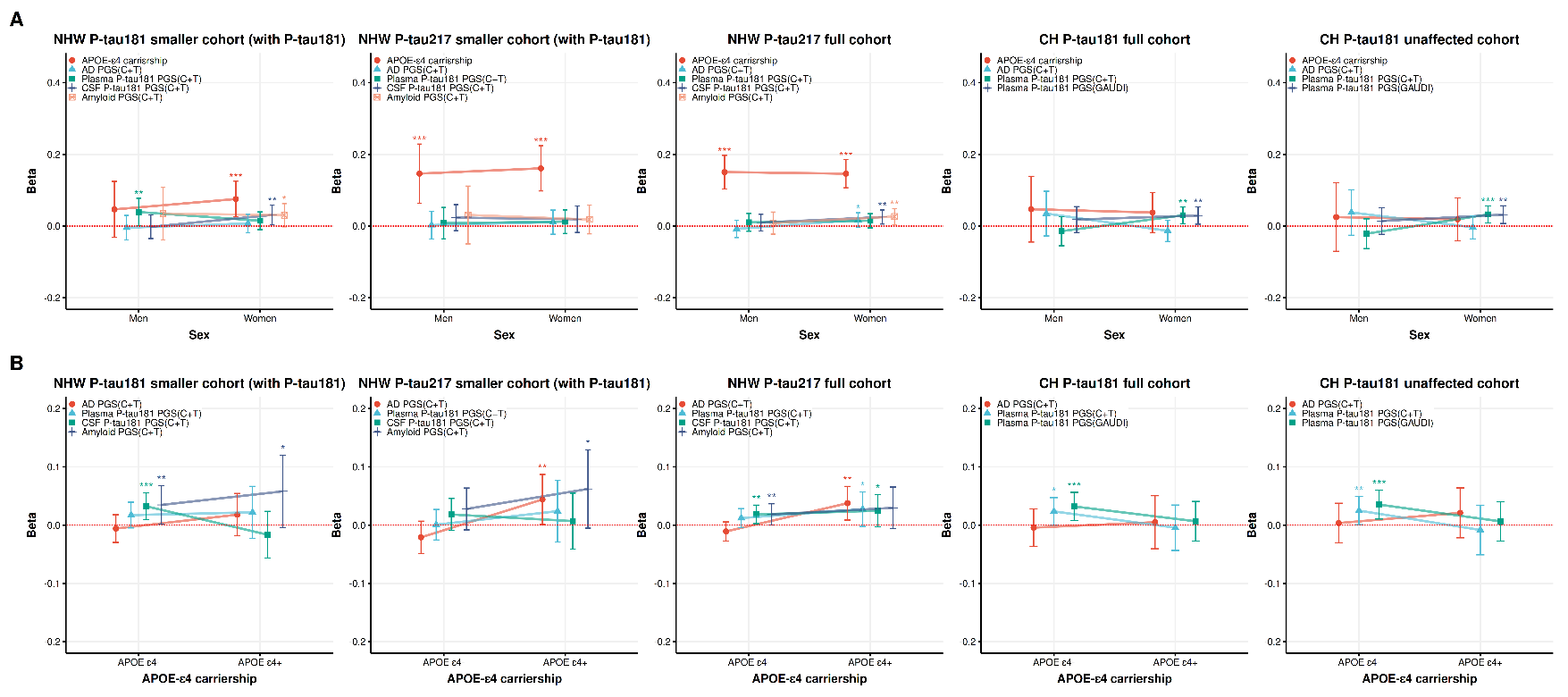

**Supplementary Figure 3** presents the sex- and *APOE* ε4-stratified longitudinal associations of *APOE* ε4 and PGSs derived from GWAS of Core-1 biomarkers and clinical AD with plasma P-tau181 and P-tau217 among White and Hispanic individuals. Panel A shows stratified analyses by sex in both non-Hispanic White and Caribbean Hispanic individuals. Panel B presents the same analyses stratified by *APOE* ε4 status. All PGS predictors were first tuned in a designated tuning sample, and the PGSs that showed the best predictive performance in that sample were applied to the target samples. For all longitudinal analyses, we used linear mixed-effects models adjusted for age, sex, and the first 10 principal components, and included random intercepts for subjects to account for within-individual correlations. For analyses involving the PGS derived from amyloid PET imaging, we excluded 89 WRAP participants who were included in the original GWAS to avoid sample overlap. Similarly, for analyses involving the PGS derived from the AD GWAS in the Hispanic sample, we excluded 219 WHICAP participants who were also included in the original GWAS. All continuous genetic predictors were z-standardized. ***p < 0.01, **p < 0.05, *p < 0.1. C+T: clumping and thresholding; GAUDI: a powerful tool for PGS analysis in admixed populations, as it explicitly models ancestry-differential effects while borrowing information across segments with shared ancestry; NHW: non-Hispanic Whites; CH: Caribbean Hispanics.

**Supplementary Figure 4. Associations Of Genetic Predictors With Longitudinal Plasma P-Tau217 Among Non-Hispanic Whites In The Wisconsin ADRC, and Stratified By Age, Sex, And *APOE* ε4 Status (N_max_ = 247, N_max_ = 299)**
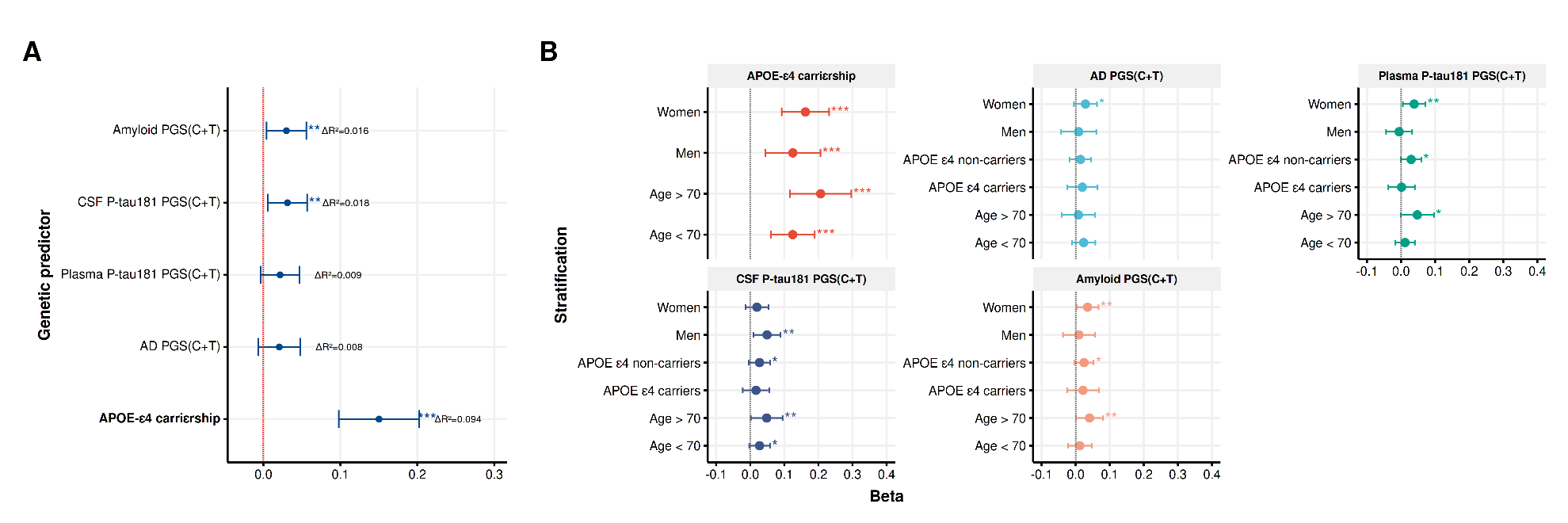

**Supplementary Figure 4** presents the longitudinal associations of *APOE* ε4, Core-1 biomarker-derived PGSs, and the AD GWAS PGS with plasma P-tau217 among non-demented, non-Hispanic White individuals in the Wisconsin ADRC (Panel A), and stratified by age, sex, and *APOE* ε4 status (Panel B). For the age-stratified analyses, participants were grouped as <70 or >70 years old due to the small sample size. All longitudinal analyses were conducted using linear mixed-effects models adjusted for age, sex, and the first 10 principal components. Random intercepts were included to account for within-individual correlations. All continuous genetic predictors were z-standardized. ***p < 0.01, **p < 0.05, *p < 0.1. C+T: clumping and thresholding; n_max_: Maximum number of unique individuals included in the sample. N_max_: Maximum number of person-level observations (i.e., person-years).

**Supplementary Figure 5. Age-, Sex-, and *APOE* ε4-Stratified Longitudinal Associations of Core-1 Integrative PGS with Plasma P-Tau217 Among Non-Hispanic White Individuals (***See main text for sample-specific sizes)*

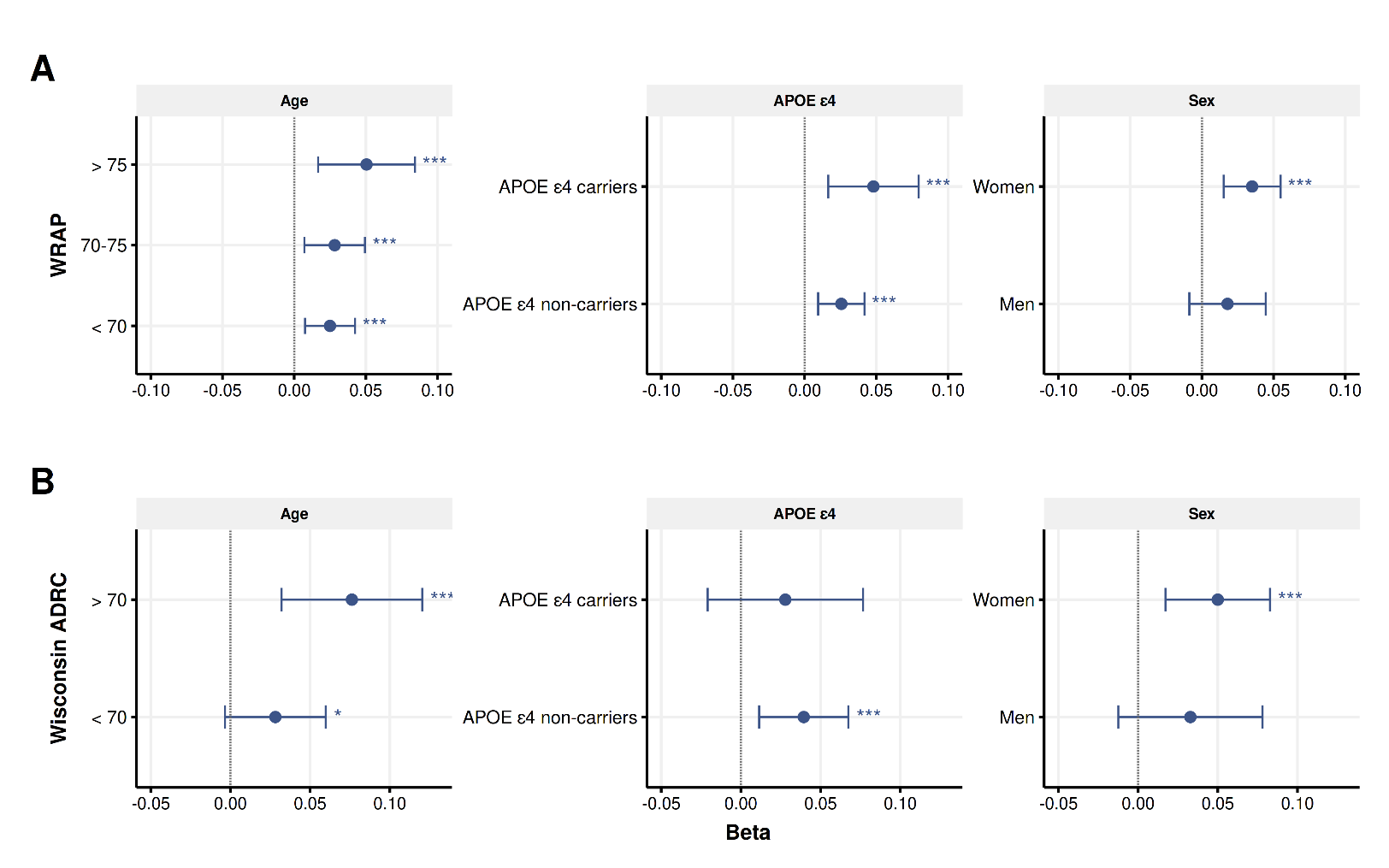

**Supplementary Figure 5** presents the age-, sex-, and *APOE* ε4-stratified longitudinal associations of the Core-1 biomarker integrative PGS with plasma P-tau217 among non-Hispanic White individuals. Panel A shows the stratified analyses in WRAP, while Panel B presents the same analyses for the Wisconsin ADRC. The integrative Core-1 biomarker PGS was an unweighted composite of three individual Core-1 PGSs, including PGSs for amyloid, CSF P-tau181, and plasma P-tau181. All individual PGSs were standardized before inclusion in the composite score. For all longitudinal analyses, we used linear mixed-effects models adjusted for age, sex, and the first 10 principal components, and included random intercepts for subjects to account for within-individual correlations. In WRAP, we excluded 89 participants who were included in the original amyloid PET GWAS to avoid sample overlap. The final integrative PGS was z-standardized. ***p < 0.01, **p < 0.05, *p < 0.1.
